# Computational Assessment of Memory Function in Kidney Transplant Recipients and Donors

**DOI:** 10.1101/2025.10.01.25337071

**Authors:** Thomas J. Wilschut, Tim J. Knobbe, Andrea Stocco, Holly Hake, Anna M. Posthumus, Aaltje L. Ziengs, TransplantLines Investigators, Hans Blokzijl, Kevin Damman, Vincent E. de Meijer, Martin H. de Borst, Anne M. Buunk, Jacoba M. Spikman, Hedderik van Rijn, Stephan J.L. Bakker

**Author notes:** Correspondence may be addressed to: prof. dr. Hedderik van Rijn, MemoryLab Health BV, Stationsstraat 10, 9711AS, Groningen, The Netherlands. Note: indicate shared first and last authorship (Thomas J. Wilschut, Tim J. Knobbe). Note: indicate shared first and last authorship (Hedderik van Rijn, Stephan J.L. Bakker).

## Abstract

Impaired memory function is a frequent yet understudied symptom in kidney transplant recipients. Here, we demonstrate a remote, minimally burdensome memory screener, the Seattle-Groningen Memory Assessment (SGMA), which estimates a patient’s speed of forgetting from paired-associate learning. We show that the memory score derived from an 8-minute session is a reliable and accurate measure of an individual’s ability for long-term retention. Kidney transplant recipients (n=556) showed more forgetting than (potential) donors (n=408). Memory scores were sensitive to demographics (age and education level), and related to self-reported sleep quality, fatigue, and health-related quality of life. On the physiological level, we were able to link more forgetting in recipients to more monocytes, neutrophils, reticulocytes, and a higher white blood cell count, as well as lower ferritin and more iron deficiency. Overall, this work highlights the potential of computational memory assessment as a minimally burdensome and reliable tool for detecting cognitive impairments in complex multimorbidity populations. The approach may be particularly valuable in research settings where detecting subtle changes in cognitive health—often missed by existing assessments—is crucial for understanding disease progression and treatment effects.

## 1 Introduction

Cognitive difficulties are a common yet understudied problem in patients with kidney failure and in kidney transplant recipients (KTR) [e.g., see 1, 2]. An increasing number of studies report deficits in memory, processing speed, and executive function [3–5], with recent evidence indicating that mild cognitive impairment—a key precursor to Alzheimer’s Disease—affects 16% of outpatient KTR [2]. Research suggests that memory deficits often persist, even after successful kidney transplantation [6]. Moreover, studies report significantly elevated risk of neurodegenerative diseases, including Alzheimer’s disease and vascular dementia, in patients with advanced kidney disease [7–9].

Accurate and accessible screening of cognitive- and memory function is valuable for both clinical practice and for sci-entific research into the mechanisms that underlie impaired cognitive function in KTR. In practice, however, widely used screening tools like the Mini-Mental State Examination (MMSE) [10] and Montreal Cognitive Assessment (MoCA) [11] face some limitations. First, the specificity and sensitivity of these tests to detect abnormalities in memory performance is generally low [e.g., see 12]. Second, their fixed, non-adaptive format may negatively affect the test-taker experience, as it may be overwhelming for patients with cognitive impairments and tedious for individuals without impairments. Third, these tests generally show large practice or test-retest effects that limit repeatability and the ability to track changes in cognitive performance over time [13].

These methodological shortcomings have tangible consequences for both clinical practice and fundamental research. In clinical practice, quick and accessible screening of cognitive impairment is valuable for assessing the need for further neuropsychological investigation, which in turn could lead to further tailored interventions—such as providing targeted support for medication adherence in patients with memory impairment or offering early cognitive rehabilitation [e.g., see 14]. For fundamental research, the lack of sensitive and repeatable memory assessments limits the ability to track cognitive trajectories over time and, as a consequence, to identify underlying neural or systemic contributors to cognitive decline. In addition, without reliable tools to measure subtle cognitive changes, it is challenging to evaluate the efficacy of targeted interventions.

To address this gap, we use a newly developed digital memory test, the Seattle-Groningen Memory Assessment (SGMA). In this memory test, participants are interleavedly asked to memorize cue-answer pairs and quizzed on their ability to recall the answer when just presented with the cue. This type of testing resembles toponym or vocabulary flashcard learning, and can cover any domain of interest to the participants. Designed for remote administration and completed in just 8 minutes, SGMA aims to provide reliable and precise assessments of long-term memory function. The assessment is adaptive and tailors the test to be optimally efficient for each participant, making it minimally burdensome. Internally, SGMA relies on computational phenotyping: it uses the patient’s data (accuracy and response times) to fit an established computational model of memory retrieval originally proposed by Anderson and Schooler [15] and Pavlik and Anderson [16]. The model simulates the accessibility or ‘activation’ of information in the patient’s declarative memory over time. Critically, this approach isolates a central measure of memory function (the speed at which information is forgotten in long-term memory) from other peripheral factors (such as perceptual and motor delays). At the end of the test, a patient’s memory capacity can be summarized in a single idiographic SGMA score, reflecting the average rate at which memory activation declined during the session. The within-session forgetting rates captured by the SGMA score generalize to memory decay over extended periods [17]. As the SGMA score can be calculated based on different item sets and reflects the *process* of memorization instead of the outcome, it avoids practice or test-retest effects and minimizes the influence of participant’s prior knowledge on the test materials [see 18].

In earlier work, the SGMA score has been shown to outperform other cognitive screening outcomes, such as General Cognitive Capacity (GCA, a measure similar to a general intelligence or IQ score) and Working Memory Capacity (WMC), in predicting long-term memory recall [19]. Neuroimaging studies indicate that the SGMA score is linked to distributed resting-state activity in EEG and fMRI data [20, 21], which is known to capture stable individual differences in brain and cognitive function. Recent clinical research has demonstrated robust correlations between the SGMA score and existing assessments memory and cognitive function, including the Consortium to Establish a Registry for Alzheimer’s Disease (CERAD) and Montreal Cognitive Assessment (MoCA) [22]. A large-scale study in collaboration with Seattle’s Alzheimer’s Disease Research Center (ADRC) and the University of Washington found that the SGMA score derived from a single 8-minute session completed at home achieved 82% accuracy in detecting mild cognitive impairment [18]. Overall, early findings suggest that the SGMA score is a fast and reliable measure of a trait-like cognitive capacity that uniquely predicts an individual’s ability for long-term retention.

In this study, we employ the SGMA to investigate memory function in KTR by conducting a cross-sectional, online behavioral study involving 408 KTR and 556 (potential) kidney donors (KD) from the TransplantLines Biobank and Cohort Study [23, 24, see Supplementary Figure 1]. The study has two primary aims. First, we evaluate the clinical feasibility, validity, and reliability of SGMA for large-scale memory screening. To do this, we analyzed patient response rates, SGMA’s sensitivity to demographic variables, its correlations with established neuropsychological assessments, and its associations with subjective memory ratings. Second, we explore how various pre- and post-transplant health factors relate to memory function in KTR, using the SGMA scores in combination with clinical, demographic, and biochemical data, and compared these patterns with those observed in (potential) KD. While directly relevant to the KTR population, this work will also provide insights to support the development of improved cognitive assessments for use in other complex, multimorbid populations.

## 2 Results

### 2.1 Baseline characteristics

#### 2.1.1 Response- and return rates suggest high engagement

The first aim of this project was to assess the feasibility of using SGMA as an online memory screener in KTR. All 2, 301 KTR or (potential) KD in the TransplantLines Biobank and Cohort study were invited by e-mail to complete up to three repeated, online memory assessments. Out of the invited participants, 964 completed at least one SGMA session, corresponding to a 41.89% overall response rate of (see Supplementary Figure 1). Of all participants who completed the first assessment, 77.05% completed the second assessment and 70.43% completed the third assessment, showing high engagement, low attrition rate, and willingness to return for repeated testing.

#### 2.1.2 Comparison of responders and non-responders

Sample characteristics for responders (i.e., those who completed at least one complete memory assessment versus those who did not) versus non-responders, for both recipients and (potential) donors, are listed in Supplementary Table 1. In the KTR group, we found no significant differences in terms of age (*p* = 0.773), sex (*p* = 0.179), years since transplantation (*p* = 0.941), pre-transplant dialysis or no pre-transplant dialysis (*p* = 0.081) or kidney function (estimated glomerular filtration rate (eGFR), *p* = 0.496) between responders and non-responders. Responders had more often received a kidney from a living (rather than deceased) donor than non-responders did (61.87% versus 54.99%, *p* = 0.009). In the (potential) KD group, responders were slightly older than non-responders (63.27 versus 60.58 years, *p <* 0.001), and had a lower kidney function (eGFR, 63.09 ± 13.80 versus 68.57 ± 16.92 *mL/min/*1.73*m*^2^, *p <* 0.001). We found no significant differences in sex and time since donation for those who donated. Overall, the comparison between responders and non-responders suggests that the sample was fairly representative of all KTR and KD in the TransplantLines Biobank and Cohort Study.

#### 2.1.3 Comparison of recipients and (potential) donors

To assess the memory function of KTR, we compared them to (potential) KD, who served as control group because of their typically high similarity to KTR in terms of socio-economic status and geographic location (e.g., often sharing the same household). This group included individuals who had been or were being screened for donation when this study was conducted. Table 1 shows the baseline characteristics for KTR and KD separately. In total, the sample consisted of 556 KTR and 408 (potential) KD. On average, recipients were slightly younger (58.59 ± 12.64 versus 63.27 ± 10.77 years, respectively, *p <* 0.001) and more more frequently male (60.01% versus 44.12% respectively, *p <* 0.001) than (potential) KD. Median time since donation or transplantation was higher in recipients than in donors (6.06, *IQR* = 2.50 − 11.32 versus 4.97, *IQR* = 2.22 − 7.72 years, respectively, *p <* 0.001). For recipients, the mean eGFR was 53.16 ± 18 *mL/min/*1.73*m*^2^, 62.05% of KTR received a kidney from a living donor and 55.94% underwent dialysis before transplantation.

**Table 1.** Sample characteristics for recipients and (potential) donors. Time since donation or transplantation shows median (IQR) values, the other rows show means (SD) or absolute numbers and proportions.

|  | Recipient | (Potential) donors | *p* |
| --- | --- | --- | --- |
| N | 556 | 408 |  |
| Age (years) | 58.59 (12.64) | 63.27 (10.77) | < **0.001** |
| N female | 222 (40%) | 228 (56%) | < **0.001** |
| Education |  |  |  |
| No education | 4 (1%) | 0 (0%) |  |
| Lower education | 13 (2%) | 4 (1%) |  |
| Lower vocational education | 89 (16%) | 47 (12%) |  |
| Intermediate secondary education | 157 (28%) | 110 (27%) |  |
| Intermediate vocational education | 67 (12%) | 61 (15%) |  |
| Higher secondary education | 44 (8%) | 32 (8%) |  |
| Higher vocational education | 136 (24%) | 120 (29%) |  |
| Academic education | 46 (8%) | 34 (8%) |  |
| Years since donation or transplantation | 6.06 (2.50–11.32) | 4.97 (2.22–7.72) | < **0.001** |
| N living donor | 345 (62%) |  |  |
| N pre-transplant dialysis | 311 (56%) |  |  |
| eGFR* (mL/min/1.73m<sup>2</sup>) | 53.16 (17.53) | 63.09 (13.80) | < **0.001** |

\*estimated glomerular filtration rate (creatinine).

### 2.2 Validity and reliability

#### 2.2.1 SGMA scores are sensitive to demographic variables

To assess the validity of the SGMA, we first examined its associations with demographic variables. Figure 1 shows SGMA scores by age, sex, and education level. Panel A demonstrates that SGMA scores, predictably, declined substantially with age. There was no noticeable association with sex. Panel B shows that higher education levels were generally associated with better performance on the memory test. The above results were corroborated by the results of a linear regression model (Table 2), which showed a significant negative association between age and SGMA score (*β* =− 0.69, *p <* 0.001), a significant positive association with education level (*β* = 2.34, *p <* 0.001), and no association with sex (*β* = −0.27, *p* = 0.765). These results align with previous studies linking age [25, 26] and education [27] to memory performance. Although prior research has occasionally reported sex differences in memory—for example, women are sometimes reported to outperform men on verbal memory tasks, while men may show advantages in spatial memory [28, 29]—these effects are typically small. Moreover, because the SGMA is designed to assess memory for image–term associations that are neither purely verbal nor spatial, it is not surprising that no robust sex differences in memory performance were observed in this study.

**Figure 1.**
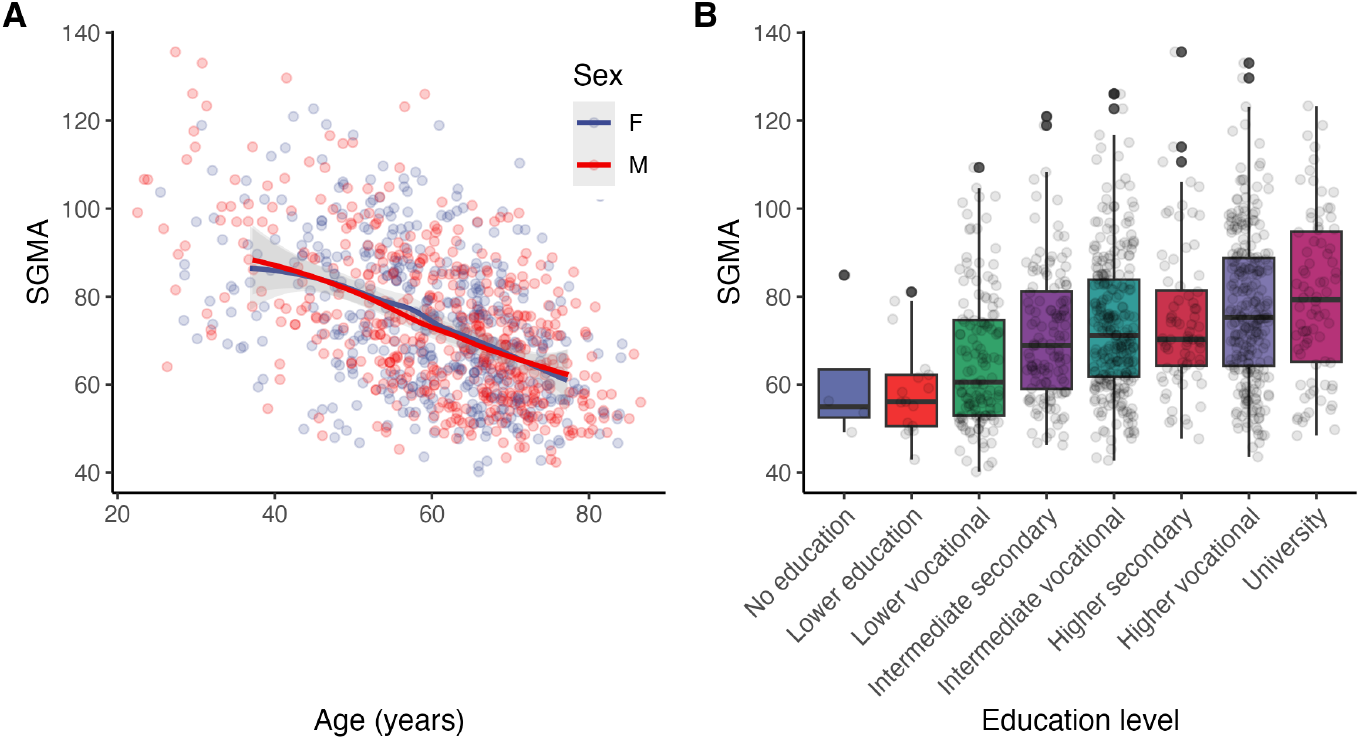
Estimated memory function (SGMA score) as a function of age, sex and education level. **Panel A** shows that the SGMA scores declined with age, but were not affected by sex. **Panel B** shows that participants with higher education levels scored better on the SGMA.

**Table 2.** Linear model for the association of age, sex, and education with the SGMA score.

|  | Estimate | Std. Error | t | p |
| --- | --- | --- | --- | --- |
| (Intercept) | 102.50 | 2.80 | 36.55 | < 0.001 |
| Age | -0.69 | 0.04 | -18.45 | < 0.001 |
| Sex (female = 0, male = 1) | -0.27 | 0.91 | -0.30 | 0.76 |
| Education level | 2.34 | 0.27 | 8.81 | < 0.001 |

#### 2.2.2 SGMA scores are reliable and show no testretest effects

For each of the three memory assessments, participants could choose their own test topic from a list of ten options (see Figure 2A) based on their personal interest. Figure 2A shows the average correlation between scores on the different topics, and panel B shows the average correlation between the SGMA scores for the three assessments. The reliability of the repeated measurements ranged between 0.61 and 0.64 over sessions, which is typically interpreted as good reliability [e.g., see 30]. Panel C shows the average estimated SGMA scores over assessments, showing that participants did not meaningfully or significantly improve with repeated testing (in fact, average performance showed a nonsignificant *decreasing* trend over assessments, with 0.91 SGMA points per session, *p* = 0.062). Overall, these results confirm that the SGMA score is a stable and repeatable measure of memory performance.

**Figure 2.**
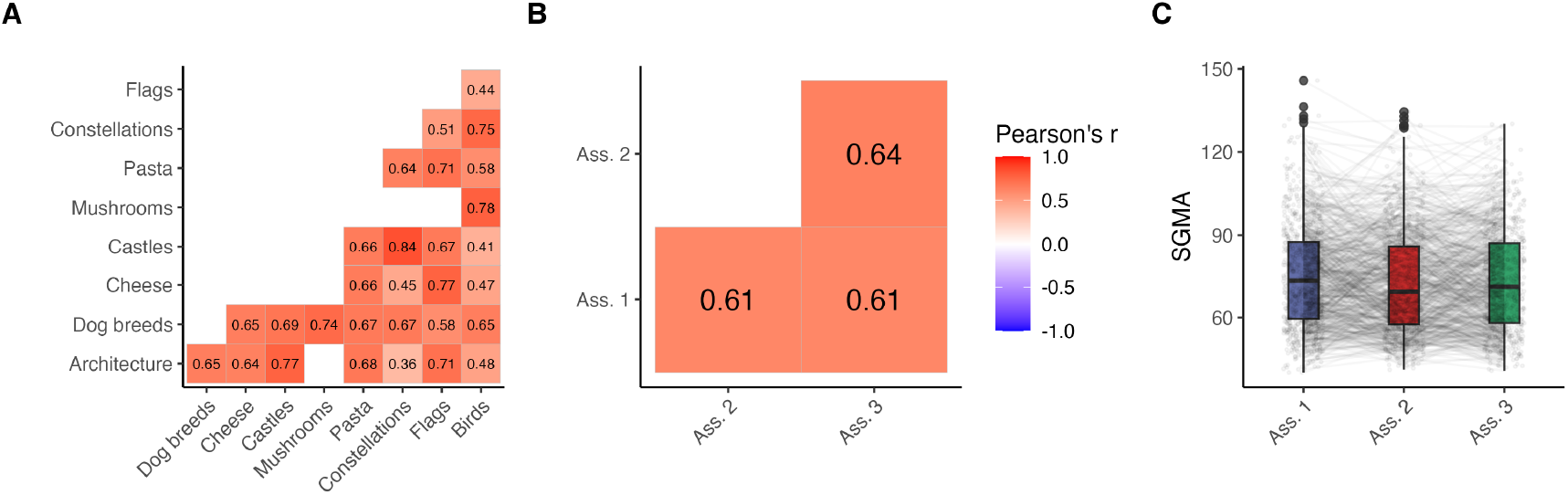
Average test-retest reliability for different test topics (**A**) and assessments (**B**). In **Panel A**, only cells for which there were 20 or more pairwise observations are shown. **Panel C** shows the SGMA score distribution over assessments, reflecting the absence of practice or test-retest effects. Lines connect observations of individual participants.

#### 2.2.3 SGMA scores are related to memory, processing speed, and attention

To determine the construct validity of the SGMA in the current clinical sample, we analyzed its associations with a battery of established neuropsy-chological assessments. Figure 3 and Table 3 show Pearson’s correlations between scores on the neuropsychological tests and the SGMA scores. Mean outcome values for neuropsy-chological tests for recipients and donors separately are provided in Supplementary Table 2.

**Figure 3.**
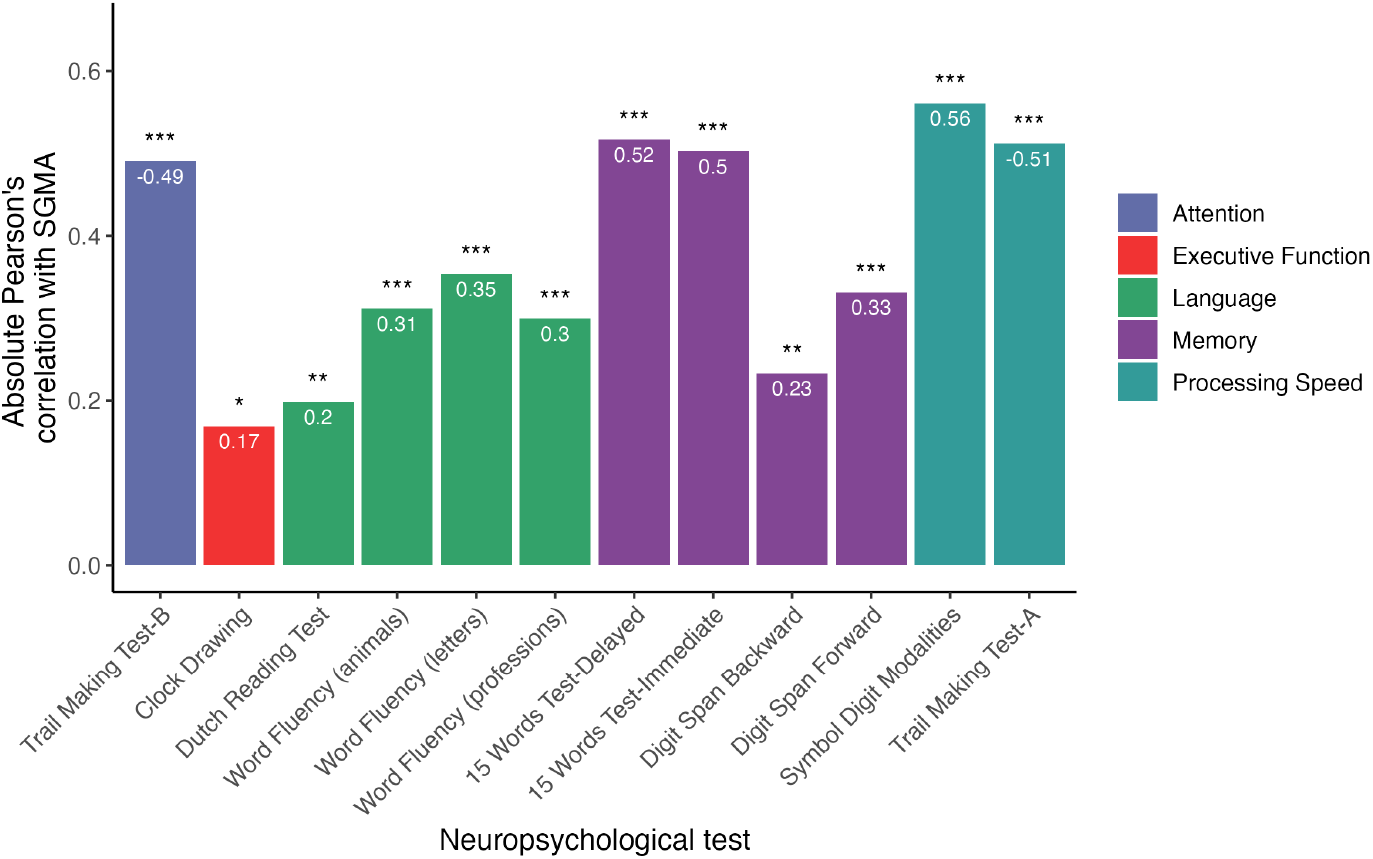
Correlations between neuropsychological tests and the SGMA score. * * *: *p* < 0.001; **: *p* < 0.01; *: *p* < 0.05.

**Table 3.**
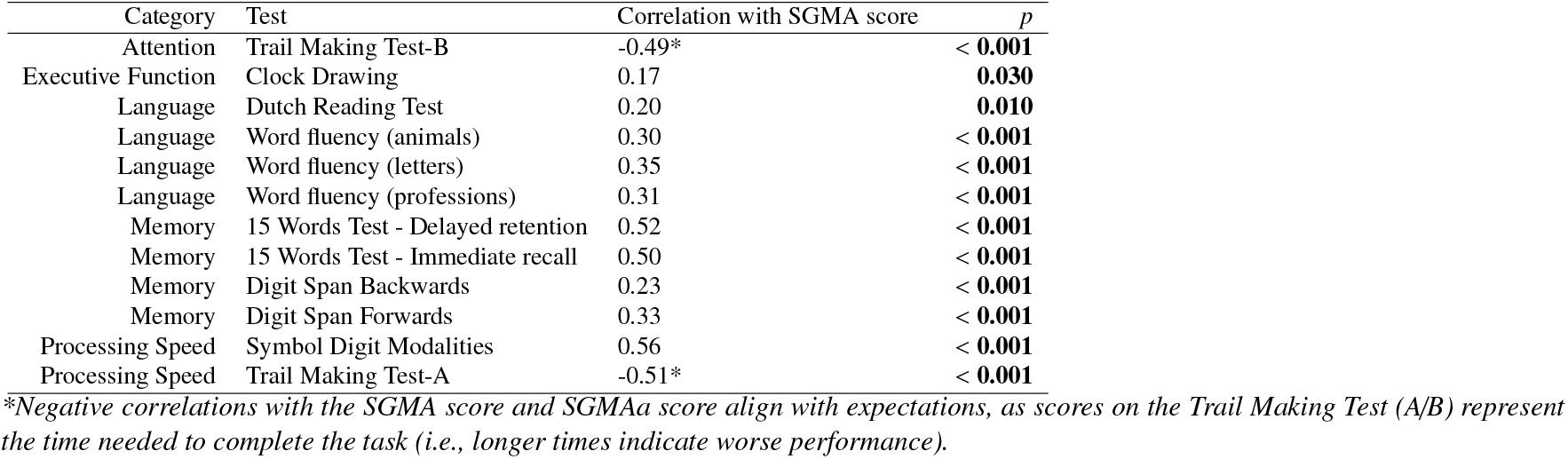
Correlations of previously performed neuropsychological tests with the SGMA score.

In line with our theoretical framework, SGMA scores showed moderate correlations with key measures of declarative memory, including both immediate and delayed recall on the 15 Words Test (where participants are presented with a list of 15 words, and must recall as many as possible immediately after each trial, and after a delay of 20 minutes). Next to associations with primary measures of declarative memory we observe moderate associations with attention (Symbol Digits Modalities Test, Trail Making Test-A), and processing speed (Trail Making Test-B): constructs that are theoretically and empirically linked to memory storage and retrieval [e.g., see 31, 32]. Correlations with language (Word Fluency, Reading) and executive functioning (Clock Drawing) tasks are notably weaker. This difference suggests that the SGMA score does not reflect general cognitive ability, but instead captures key mechanisms related to memory processes.

It is good to note that neuropsychological tests were completed several years prior to the memory assessment during a study visit for the TransplantLines Biobank and Cohort study (median time since completion = 6.26 years, *IQR* = 5.77 − 7.22 years). As it is possible that at least some of these neuropsychological measures changed in a nonlinear manner during the years between assessments, the absolute strength of the observed correlations may need to be interpreted as conservative *under*estimations of the true associations. However, Supplementary Figure shows that the strength and direction of correlations between SGMA and neuropsychological test scores were comparable across subgroups split by time since neuropsychological assessment, indicating minimal influence of assessment recency on the observed associations. This suggests that the associations with SGMA scores are robust, supporting the interpretation that the SGMA captures stable, trait-like differences in memory function.

#### 2.2.4 The accuracy of memory self-assessment declines with age

Before starting the first memory assessment, all participants were asked to provide a subjective assessment of their own every-day memory performance. Responses were provided (on a 4-point Likert scale) with options (1) ‘poor’; (2) ‘average’, (3) ‘good’; (4) ‘very good’. Table 4 shows the results of a linear regression model estimating the observed SGMA score from subjective memory rating (1–4) and age (also see Supplementary Figure 4). We found a significant association between subjective memory and the SGMA score: people are able, at least to some extent, to rate their own memory function (β = 9.94, *p* < 0.001). There is also an association with age, indicating that older people rate their memory function lower (β = −0.42, *p* < 0.001). Finally, we find an interaction between age and subjective memory, indicating that the association between age and the SGMA score is weaker for people with lower subjective memory ratings (β = −0.13, *p* < 0.006). In other words, older people made less accurate judgments about their own memory function.

**Table 4.** Linear model, estimating the SGMA score using subjective memory rating and age.

|  | Estimate | Std. Error | t | p |
| --- | --- | --- | --- | --- |
| (Intercept) | 92.51 | 7.74 | 11.94 | < 0.001 |
| Subjective memory rating | 9.94 | 2.84 | 3.50 | < 0.001 |
| Age | -0.42 | 0.12 | -3.37 | 0.001 |
| Subjective memory rating × Age | -0.13 | 0.05 | -2.78 | 0.006 |

### 2.3 Interim summary

Together, the results from the preceding sections demonstrate that SGMA is a feasible, reliable, and valid tool for assessing memory function in KTR and KD. Using SGMA resulted in high response and return rates, suggesting that it is well-received and accessible in clinical populations. The SGMA score was reliable across repeated sessions and showed no practice effects. SGMA scores were sensitive to known demographic predictors of memory performance, and showed robust correlations with established neuropsychological measures—particularly those indexing verbal memory, processing speed, and attention. Overall, by establishing the SGMA as a reliable and valid tool for remote memory assessment, they provide a foundation to examine how clinical and physiological factors contribute to memory function in KTR—the second aim of this study.

### 2.4 Associations with clinical variables

#### 2.4.1 KTR show lower memory performance than (potential) KD

To properly compare KTR and KD while accounting for demographic confounds, we created an adjusted version of the SGMA score, referred to as SGMA-adjusted (SGMAa), in which the SGMA score is adjusted for age, education and sex. This adjusted score was calculated using the residuals of the linear model shown in Table 2. We compared the memory performance of KTR with (potential) KD using the SGMAa score (thus adjusted for demographic variables); Figure 4 provides an overview of the results. Panel A shows that recipients had significantly lower SGMAa scores compared to (potential) donors (mean difference = 2.19, *p* = 0.010). This result can be interpreted as a small but statistically significant reduction in memory performance compared to healthy individuals, potentially reflecting the lasting cognitive impact of kidney failure, comorbidities, or factors related to the transplantation (e.g., immunosuppressive treatment). Panel B shows that SGMAa scores were not significantly associated with time since transplantation or donation (recipients: *p* = 0.423, donors: *p* = 0.816). In other words, these results suggest that neither kidney transplantation nor donation influences the normal age-based decline in memory performance. This suggests that the observed lower memory scores in KTR are unlikely to reflect progressive post-transplant cognitive decline, but rather reflect the impact of pre-transplant health factors^1^.

**Figure 4.**
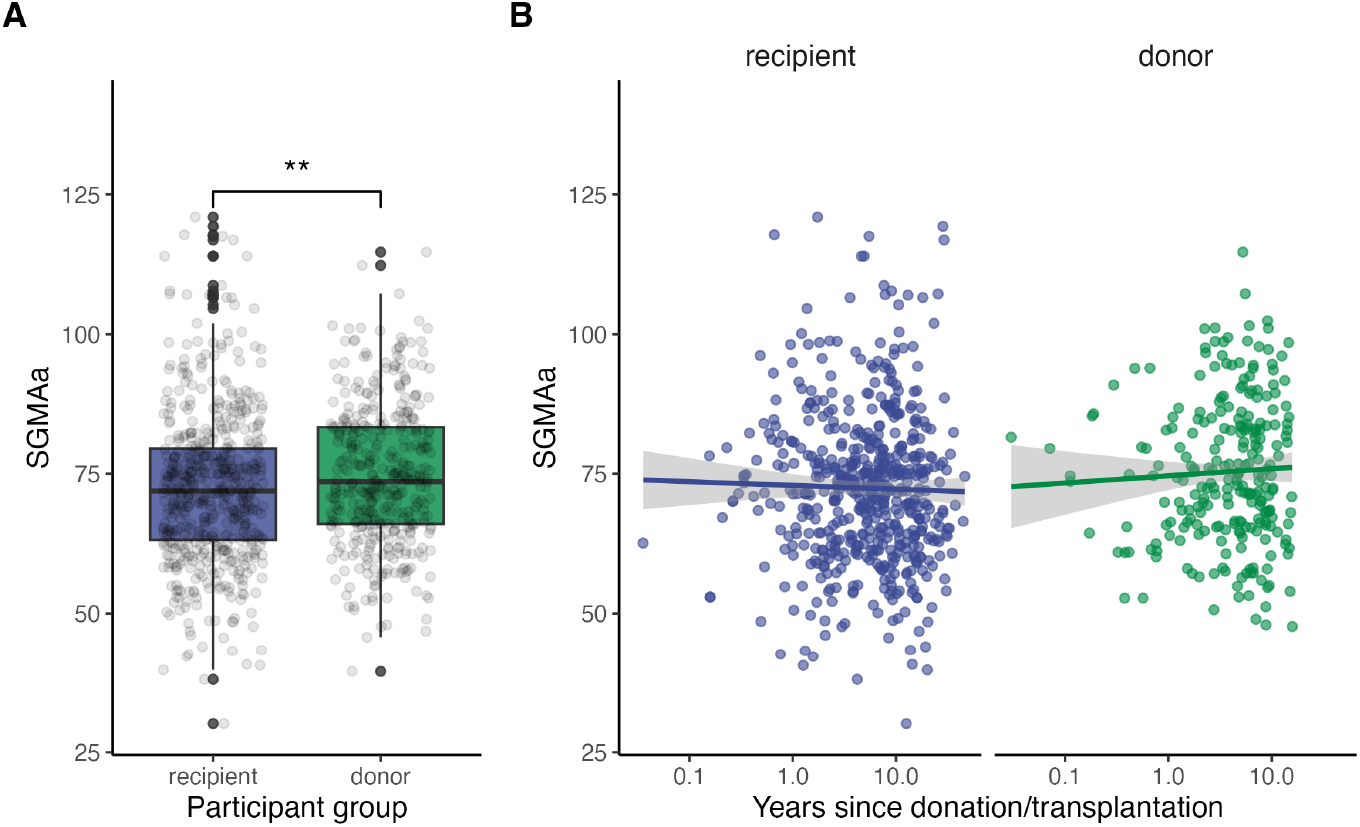
Memory performance in KTR and (potential) KD, based on SGMA scores adjusted for age, sex, and education (SGMAa). **Panel A:** KTR show significantly lower SGMAa scores compared to (potential) KD, indicating reduced memory performance in the recipient group. **Panel B:** the SGMAa scores were not significantly associated with time since transplantation or donation.

To further validate these findings, we conducted a sensitivity analysis comparing KTR with their living KD, in order to achieve better demographic comparability in terms of age, time since transplantation/donation, and geographical area (e.g., often sharing the same household). An independent-samples t-test which included 96 KTR and 96 KD, yielded results consistent with the primary analysis, with significantly lower SGMAa scores among KTR compared to their living kidney donors (*p* = 0.028), as shown in Supplementary Figure 5. This reinforces the conclusion that the observed differences in memory function are not explained by demographic confounding.

Recipients with Cystic kidney disease as etiology of their kidney failure had higher SGMAa scores than recipients with diabetic nephropathy as etiology of their kidney failure (*p* = 0.028). No differences were between recipients with other etiologies of their kidney failure were found, as presented in Supplementary Table 3 and Supplementary Figure 6. Within the recipient group, we found no significant difference in the SGMAa scores between those who had received pretransplant dialysis and those who had not (*p* = 0.867). Similarly, no significant difference was observed between recipients of a kidney from a living versus a deceased donor (*p* = 0.173). Our findings point to a broader cognitive vulnerability that persists post-transplantation. However, we note that the time since transplantation was variable across individuals, and subtle progressive effects or delayed recovery processes cannot be ruled out based on the current results.

#### 2.4.2 Immunosuppressive medication in recipients

To examine whether immunosuppressive medication use was related to memory performance. No significant differences in adjusted SGMA scores were found for calcineurin inhibitors (*p* = 0.72), mTOR inhibitor (*p* = 0.07), proliferation inhibitor (*p* = 0.76), or corticosteroid (*p* = 0.92) use. These null findings likely reflect the limited variability in medication regimens—most recipients used similar combinations of immunosuppressants—which reduces the likelihood of detecting differential cognitive effects across groups.

#### 2.4.3 Self-reported sleep quality, fatigue, and HRQoL are associated with SGMAa scores

To explore how general well-being relates to memory performance, we examined associations of self-reported measures of sleep quality, fatigue, and health-related quality of life (HRQoL), collected at the time of the third memory assessment, with SGMAa scores. Sleep quality was measured using the Pittsburgh Sleep Quality Index (PSQI), where higher scores indicate poorer sleep. We observed a modest but significant negative correlation with SGMAa (*r* = −0.13, *p* = 0.003), suggesting that individuals who reported poorer sleep tended to show lower memory performance. Fatigue was assessed with the Checklist Individual Strength Revised (CIS-8), where higher scores reflect greater fatigue. A similar pattern emerged, with higher fatigue levels associated with lower SGMAa scores (*r* = −0.09, *p* = 0.017). Finally, we measured HRQoL using the Short Form 12 (SF-12). Here, we found a positive correlation with the SGMAa for the physical component score (*r* = 0.17, *p* < 0.001), indicating that individuals who perceived their overall physical health and functioning more positively performed better on the memory test. We also found a trend in the direction of an association for the mental component score, with higher scores related to better memory performance (*r* = 0.07, *p* = 0.054). Although the above-mentioned associations are relatively small, they are consistent for reported sleep quality, fatigue, and HRQoL. Together, they suggest that broader aspects of health and daily functioning are related with memory function as captured by the SGMA, also when partialling out potential effects of demographic factors.

### 2.5 Physiological markers

We analyzed the associations between commonly used biomarkers, collected either during routine clinical care or at the last TransplantLines study visit (see section 4.1), and memory function to gain insight into long-term physiological processes that may affect memory function.

#### 2.5.1 Blood markers explain variance in memory scores

To assess the overall explanatory power of blood markers on memory function, we conducted four separate analyses. Given the large number of collected blood markers, we used LASSO-regression models, which apply an *L*_1_ regularization penalty to mitigate overfitting, handle multicollinearity, and perform variable selection by shrinking less informative predictors toward zero [33]. Models were conducted separately to predict (a) unadjusted SGMA scores in recipients, (b) adjusted SGMAa scores in recipients, (c) unadjusted SGMA scores in donors, and (d) adjusted SGMAa scores in donors. The left panel of Figure 5 shows the standardized coefficients from the surviving regressors from each of these models. Remarkably, blood markers alone accounted for 36.66% and 25.52% of the variance in unadjusted SGMA scores in KTR and KD, respectively. After demographic adjustment, the variance explained remained substantial at 9.97% in recipients, but dropped to just 1.11% in donors. This demonstrates that—while the relationship between memory and physiology is partially mediated by demographic factors—physiological markers in KTR independently explain memory performance in recipients, even after accounting for age, sex, and education. The greater explained variance in KTR compared to KD likely reflects the fact that the physiological profiles were much less variable across individuals in KD compared to KTR (see Supplementary Figure 7). In the next sections, we discuss a number of key associations of interest in more detail.

**Figure 5.**
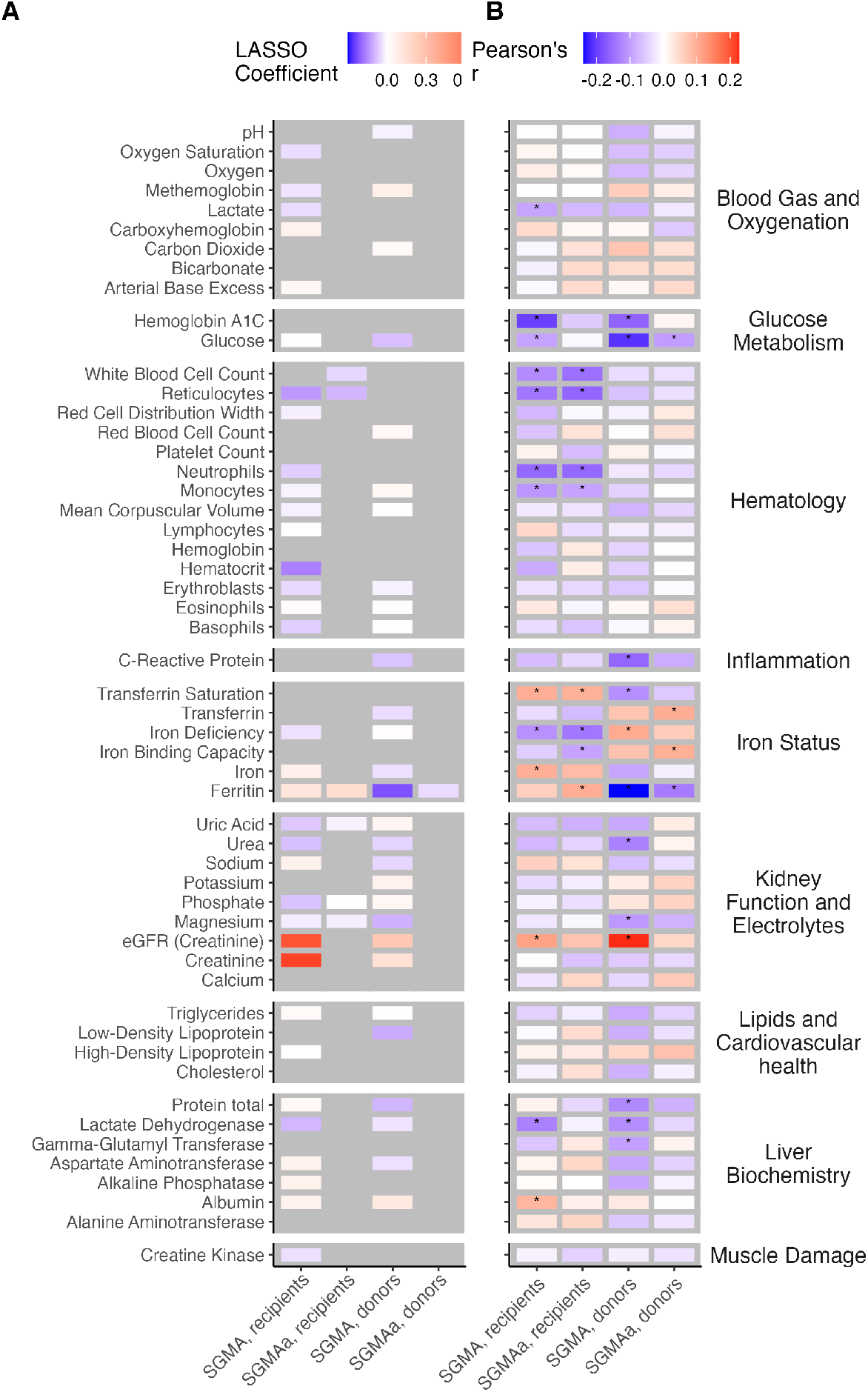
Association between sampled blood markers and memory performance. **Panel A** (left) shows LASSO regression coefficients, for models predicting the unadjusted SGMA score for recipients, the SGMAa score for recipients, the unadjusted SGMA score for donors and the SGMAa score for donors using standardized predictors. Predictors excluded from the penalized regression (coefficients of 0) are uncolored (gray). **Panel B** (right) shows individual Pearson’s correlation coefficients between measurements (vertical axis) and outcome measures (horizontal axis, from left to right: the unadjusted SGMA score for recipients, the SGMAa score for recipients, the unadjusted SGMA score for donors, and the SGMAa score for donors. Stars indicate significant correlations at the level of *p* < 0.05.

#### 2.5.2 Biomarkers and memory function in recipients

Several key associations between blood markers and memory scores emerged among recipients, as visualized in the bottom panel of Figure 5. For readability, we only report correlations based on the demographically adjusted scores (SGMAa) here, unless these were not statistically significant, in which case the unadjusted SGMA scores are explicitly reported. A comprehensive overview is available in Supplementary Table 4.

##### Glucose metabolism

Both glucose (*r* = −0.09, *p* = 0.039) and glycated hemoglobin (HbA1c, *r* = −0.20, *p* < 0.001) were negatively correlated only with the unadjusted SGMA scores, aligning with known physiological and socioeconomic demographic influences on glucose metabolism (For example, aging is linked to reduced insulin sensitivity and β-cell function [34], while sex differences in hormones and body composition affect glucose regulation [35]; lower educational attainment is also associated with adverse health behaviors that are related to poorer glycemic control [36]). We did not find evidence for a link between memory function and glucose metabolism independent of these demographic mediators.

##### Hematology

Higher neutrophil (*r* = −0.16, *p* = 0.001), monocyte (*r* = −0.09, *p* = 0.044), and white blood cell counts (*r* = −0.15, *p* = 0.001) were associated with lower SGMAa scores, suggesting that (subclinical) immune activity may be linked to reduced cognitive performance, even after demographic adjustment. Although reticulocyte count was negatively associated with SGMAa (*r* = −0.16, *p* = 0.001), hemoglobin was not associated with SGMA(a). An explanation is that the association with hemoglobin only emerges below a certain threshold, as studies showed that anemia is associated with cognitive impairment [37]. Elevated reticulocyte count may indicate active hematopoietic stress—such as chronic blood loss, inflammation, or increased red blood cell turnover, which could cause lower memory function.

##### Iron status

Ferritin (*r* = 0.10, *p* = 0.026) and transferrin saturation were positively correlated with the SGMAa scores (*r* = −0.10, *p* = 0.034). Iron deficiency and iron binding capacity (*r* = −0.10, *p* = 0.041) negatively correlated with the SGMAa scores (*r* = −0.14, *p* = 0.013). Total iron levels were only significantly associated with the unadjusted SGMA scores (*r* = 0.10, *p* = 0.029; SGMAa scores: *p* = 0.066). These findings suggest that indicators of poor iron status may be linked to reduced memory performance—even after adjusting for demographic factors. This is consistent with a recent study among KTR, in which iron deficiency was associated with impaired neurocognitive tasks measuring memory, mental speed, and attention and executive functioning [38].

##### Kidney function

eGFR correlated positively with the unadjusted SGMA scores (*r* = 0.11, *p* = 0.008), but this association disappeared after demographic adjustment. This attenuation suggests that the observed relationship was likely driven by age-, sex- or education-related differences in kidney function, rather than reflecting a direct effect of kidney function on memory performance [39]. An alternative explanation may be that by correcting for age, differences in cognitive function caused by premature decline as a result of kidney failure are adjusted for in the SGMAa, but are shown in the unadjusted SGMA.

Together, the above results demonstrate that in KTR, a substantial proportion of variance in memory performance can be explained by blood-based biomarkers alone. Notably, even after adjusting for demographic factors such as age, sex, and education, meaningful associations persist, particularly in domains related to hematologic profiles and iron status.

#### 2.5.3 Biomarkers and memory function in (potential) donors

In line with the LASSO regression results, fewer significant associations were observed among (potential) donors, likely reflecting their more stable physiological status compared to recipients. Still, some notable patterns emerged, as described below.

##### Glucose metabolism

Higher glucose levels negatively correlated with the SGMAa scores (*r* = −0.10, *p* = 0.041).

##### Hematology and inflammation

C-reactive protein negatively correlated with the SGMA scores (*r* =− 0.16, *p* = 0.001), though this association weakened after demographic adjustment (*r* =− 0.09, *p* = 0.083). Unlike in recipients, we found no associations with monocytes, neutrophils, reticulocytes, or white blood cell counts.

##### Iron status

Ferritin was negatively associated with SGMAa scores (*r* = −0.14, *p* = 0.006), while iron binding capacity showed a positive association (*r* = 0.10, *p* = 0.045). Iron deficiency positively correlated only with the unadjusted SGMA score (*r* = 0.13, *p* = 0.036), though this result should be interpreted with caution, as only 10 donors were classified as iron-deficient (see section 4.6). Transferrin showed a negative correlation with SGMAa scores (*r* = −0.10, *p* = 0.040), transferrin saturation was negatively associated only with the unadjusted SGMA score (*r* = −0.12, *p* = 0.018). Notably, this pattern is opposite to what we observed in KTR, and reflect a complex, possibly *protective* role of mild iron deficiency in (potential) donors, aligning with prior evidence linking mild iron depletion to reduced cardiovascular risk [40] and decreased risk of neurodegenerative diseases [41]. The divergence from the results found in KTR is not entirely surprising: in KTR iron deficiency often co-occurs with underlying disease activity or chronic inflammation. In such cases, iron-related markers can reflect ongoing systemic processes, and the typical associations may “flip”—a phenomenon commonly seen when iron homeostasis is disrupted in the context of illness.

##### Kidney function and liver biochemistry

Several associations initially significant with the unadjusted SGMA scores (eGFR: *r* = 0.23, *p* < 0.001; urea: *r* = −0.13, *p* = 0.008; magnesium: *r* = −0.11, *p* = 0.033; gamma-GT: *r* = − 0.10, *p* = 0.047; LDH: *r* = −0.12, *p* = 0.017) lost significance after demographic adjustment.

The fewer and weaker associations observed in KD compared to KTR are both expected and informative, reflecting the donors’ generally healthier and more stable physiological profiles. In addition, several patterns (e.g., associations with glucose, glycated hemoglobin, and eGFR, although mediated by age, sex, or education level) mirrored the associations found in recipients. This consistency between groups strengthens confidence in the relevance of these physiological factors for memory performance, even in populations with minimal clinical burden.

An important consideration in interpreting these results is the temporal gap between the laboratory measurements and the memory assessments. In our sample, the median interval was 2.28 years (*IQR* = 0.73̆4.72 years), meaning that physiological parameters and cognitive outcomes were not always assessed concurrently. Supplementary Figure 8 shows that, when stratifying the sample by time since laboratory assessment, correlation patterns between blood markers and SGMA scores were broadly consistent across subgroups. Nonetheless, replication of these analyses with *concurrent* laboratory and SGMA data would be valuable, as it would allow for more precise estimation of short-term physiological influences on memory performance and reduce the potential for temporal confounding.

## 3 Discussion

This study had two primary objectives: first, to assess the feasibility, reliability, and validity of the SGMA as a memory screening tool in a clinical population of KTR; and second, to use the SGMA scores to study how pre- and post-transplant health factors relate to memory function in KTR.

### 3.1 The SGMA as a feasible tool for memory assessment in KTR

A practical cognitive screening tool should be accessible (low-burden, time-efficient, and suitable for remote use), reliable (yielding stable results over repeated measures), and valid (consistently capturing a distinct cognitive construct). Our findings suggest that the SGMA meets these criteria in a clinical context.

First, the SGMA demonstrated excellent viability. Response- and return rates were high, especially after the initial session, suggesting that participants were engaged and the test itself was well-received. In the current project, even individuals affected by complex chronic health conditions were able to complete repeated memory assessments using the remote digital platform.

Second, the SGMA scores demonstrated good test-retest reliability, and memory scores remained stable across three repeated assessments, with no evidence of test-retest effects. This makes the SGMA well-suited for tracking cognitive change longitudinally.

Third, the SGMA scores were correlated with established neuropsychological measures of memory, attention, and processing speed, despite these having been administered a median of 6.26 years earlier. Although the temporal gap between the evaluations is a methodological limitation, the presence of significant associations despite this delay suggests that the SGMA captures stable individual differences in cognitive function. Given the individual nature of cognitive and health decline over time, the observed correlations likely underestimate the true validity that SGMA scores would show with concurrent benchmarks.

Given the above, the SGMA addresses several key limitations of conventional cognitive screening tools (e.g., MoCA, MMSE). While such instruments are widely adopted in clinical settings, their nonadaptive nature often has a negative impact on the test-taker experience. Moreover, substantial test–retest effects limit the reliability of these screening tools in longitudinal contexts [13]. The SGMA accommodates these challenges by offering an accessible alternative that maintains reliability and validity, making it well-suited for repeated cognitive screening in clinical settings. It is good to note that successful completion of the SGMA (like the MoCA, CERAD, and MMSE) does presuppose basic literacy, which may limit its applicability in some populations.

### 3.2 Memory function in KTR

In the second part of this study, our objective was to evaluate the memory function in KTR by comparing their performance to that of (potential) KD and examining associations with clinical variables, including (pre)transplant factors and time since surgery.

KTR showed significantly lower memory performance compared to KD, even after adjustment for demographics. Within KTR, memory scores were not associated with time since transplantation, nor with dialysis history or donor type. These findings are consistent with previous results from the TransplantLines Biobank and Cohort study, where 16% of KTR met the criteria for mild cognitive impairment, compared with 2.6% of KD, based on neuropsychological testing. Consistent with our current findings Ziengs et al. [2] likewise reported no association of cognitive outcomes with time since transplantation, dialysis history, or donor type. These findings suggest that cognitive deficits in recipients are persistent after transplantation and may reflect broader, lasting effects of kidney failure and/or its treatment, which is consistent with the results of[6]. Further research is needed to explore the underlying mechanisms driving these differences, including the potential impact of immunosuppressive medication [42–44, e.g., see], comorbidities and the clinical course early post-transplantation.

Our analysis of blood markers provided additional insight into the physiological correlates of memory function. Among recipients, lower eGFR, elevated reticulocyte counts, higher ferritin, glucose, and glycated hemoglobin levels, as well as increased monocyte and neutrophil counts, were associated with poorer memory performance. While some associations were attenuated after adjusting for demographic variables, several remained robust, particularly those related to iron status and hematology. Among (potential) donors, the number and strength of the associations were generally lower, consistent with the donors’ more stable health status and narrower physiological variability. It is important to note that the cross-sectional nature of the study precludes any conclusions about the directionality or causality of the observed associations between physiological markers and memory performance. Additionally, blood samples and SGMA sessions were not completed concurrently; the median interval between these measurements was 2.26 years. This temporal gap likely attenuated the strength of the actual associations, suggesting that the true relationships between physiological health and cognitive function may be more robust when measured simultaneously. The same limitation applies to the neuropsychological test battery data, which was also administered years before the memory test. Although strong associations were observed, and analyses of time since neuropsychological test or blood measurement (see Supplementary Figures 3 and 8) point to relatively robust results regardless of the recency of the measurement, future research should be conducted with concurrent tests to further validate these results.

### 3.3 Future directions and implications

Future research should employ a longitudinal design with repeated SGMA sessions and concurrent biomarker collection to more accurately study temporal dynamics and potentially causal pathways linking systemic health to cognitive change. SGMA uniquely allows for this requirement of repeated testing.

This study demonstrates the potential of computational memory assessment as a feasible, accurate, and scalable tool for cognitive screening in complex clinical populations. The SGMA combines a patient-friendly design with good psychometric properties and sensitivity to physiological burden, making it ideal for both scientific research and clinical monitoring.

In clinical settings, the SGMA offers significant opportunities for scalable cognitive monitoring, particularly in populations at elevated risk of cognitive impairment. KTR are known to face increased risk of developing mild cognitive impairment and Alzheimer’s disease, yet cognitive assessment is not routinely implemented in their standard care. Our findings demonstrate that the SGMA is not only feasible in this population but also shows good reliability and validity. These properties position the SGMA as a viable tool for regular, remote monitoring of cognitive status. The results of this screening could lead to further neuropsychological investigation and subsequently facilitate earlier identification of decline and support targeted interventions such as cognitive rehabilitation or medication adherence support. More broadly, our results suggest that the SGMA captures clinically meaningful variance in cognitive function, even after demographic adjustment, reinforcing its potential role in personalized care strategies and long-term outcome monitoring in individuals with complex health conditions.

## 4 Methods

### 4.1 Participants

An invitation to complete the three SGMA sessions was sent to all enrolled and alive participants of the TransplantLines Biobank and Cohort study [23]. This study, initiated in June 2015, invited all (potential) solid organ transplant recipients and donors (≥ 18 years old) from the University Medical Center Groningen (UMCG) to participate (participation rate of 80%). Written informed consent was obtained before inclusion. The TransplantLines study protocol was approved by local Institutional Review Board (METc 2014/077), adheres to the UMCG Biobank Regulation, and is in accordance with the WMA Declaration of Helsinki and the Declaration of Istanbul [23]. The study protocol has been described previously (see Eisenga et al. [23] and Posthumus et al. [24]).

For the current study, we included all KTR or (potential) KD who completed at least one complete SGMA session that fell outside of the exclusion criteria (see section 4.3). This resulted in a sample of 556 KTR and 408 (potential) KD (see section 2.1).

### 4.2 Procedure

Eligible participants of the TransplantLines Biobank and Cohort Study (see section 4.1) were invited using the REDCap survey and database managing software [45]. In total, participants were invited to up to three measurement sessions via email. The first invitations to participate were sent on July 17th, 2024. Before starting the first memory measurement (see section 4.3), participants were asked to complete three questionnaires on general well-being (see section 4.5). Exactly 7 days after completion of the first memory test, participants received an invitation to participate in the second memory test. Likewise, exactly 7 days after completion of the second memory test, an invitation for the third test followed. Participants who did not complete the test received a first reminder three days after each invitation, and a second reminder three days after that. A timeline of the most important events in this study is summarized in Supplementary Figure 2.

### 4.3 Seattle-Groningen Memory Assessment

Participants received invitations to complete three digital SGMA sessions, spaced one week apart. Participants were able to choose from an array of test topics based on personal preference, including Architecture, Dog breeds, Cheeses, Castles, Mushrooms, Pasta, Constellations, Flags, and Sailing Knots (participants could not choose the same topic for multiple tests). SGMA sessions took 8 minutes to complete, and within each session, items were adaptively introduced based on the performance of the participants. For each item, response latencies and accuracy scores were recorded and used to estimate a *speed of forgetting* parameter, which was continuously updated throughout the session. The final speed of forgetting parameter estimated at the end of the session, averaged over all items and sessions, was converted to an individual SGMA score using a sigmoid function (for more details on the adaptive item scheduling algorithm and the estimation of the speed of forgetting from responses, see Hake, Velde, Leonard, Grabowski, Rijn, and Stocco [18] and Sense, Behrens, Meijer, and Rijn [46].

SGMA sessions were screened for irregularities to ensure high data quality, particularly given the online nature of data collection. We applied relatively strict exclusion criteria for this purpose. Sessions that included a pause exceeding 60 seconds were excluded, as long interruptions or distractions during the task compromise the accurate estimation of forgetting speed over time. Similarly, sessions that were terminated after less than 6 minutes were removed, as they reflect incomplete data or disengagement. We also excluded sessions with an average accuracy below 50.00%, and sessions with a median response time above 8 seconds, which may indicate non-compliance, a lack of understanding of the task, or extreme inattentiveness. In total, 11.80% of all trials were excluded based on these above criteria.

To further ensure comparability across individuals and sessions, all SGMA scores were adjusted for average topic difficulty, using the residuals of a linear regression model estimating SGMA score from test topic.

### 4.4 Neuropsychological tests

Between June 2015 and March 2021, participants of TransplantLines were randomized to undergo either an extensive physical or neuropsychological assessment. Consequently, a subset of the participants who completed the SGMA had completed a series of neuropsychological tests during this period. These tests were administered by a trained researcher with a background in psychology and typically took between 30 and 45 minutes to complete. The tests were conducted a median of 6.26 years (*IQR* = 5.77 − 7.22 years) prior to completion of the first memory test. The following constructs were included in the assessment:

#### Attention

The Trail Making Test part B (**Trail Making Test-B**) assesses attention, cognitive flexibility, executive functioning, and task-switching ability. Participants are asked to connect alternating sequences between numbers and letters in ascending order (e.g., 1-A-2-B-3-C…). The task requires maintaining attention while switching between sequences. The completion time is recorded, with faster times indicating better performance [47].

#### Executive functioning

The **Clock Drawing** Test assesses visuospatial skills, executive function, and memory. Participants draw a clock, place numbers correctly, and set the hands to a given time. Errors in layout, sequencing, or time placement can indicate cognitive impairment. Scoring is based on accuracy, with points assigned for correct shape, number placement, and hand positioning [48].

#### Memory

The Wechsler Adult Intelligence Scale IV is an intelligence test for adults that includes working memory tests. One of these sub-tests, the Digit Span, has two parts: in the first part, the participant must repeat a sequence of digits in the same order, which tests working memory (**Digit Span Forwards**). In the second part, they repeat the digits in reverse order, resulting in a more demanding test of working memory (**Digit Span Backwards**) [49].

The 15 Words Test (**15 Words Test**) evaluates verbal memory. Participants are presented with a list of 15 unrelated words over five consecutive trials and must recall as many as possible immediately after each trial (Immediate recall). A delayed recall (Delayed retention) test is also conducted after 20 minutes [50]

*Language* **Word fluency** is a verbal task assessing language fluency and semantic memory. Participants must generate as many words as possible within a given category (words with a specific first letter (l), certain professions (p) and animals (a)) in one minute, and their total score is based on the number of words produced [e.g., see 51].

The **Dutch Reading Test** for Adults is a test used to estimate the general intelligence and cognitive function. The test consists of a series of words with irregular pronunciation, meaning they do not follow the usual pronunciation rules. The patient is required to read these words aloud. The NLV is a validated estimate of (verbal) intelligence. The score is determined by the number of correctly pronounced words [52].

#### Processing speed

Trail Making Test part A (**Trail Making Test-A**) measures mental speed and involves connecting 25 numbers in ascending order, as quickly as possible. Completion of the tests is timed in number of seconds [47].

The **Symbol Digit Modalities Test** assesses processing speed, attention, and cognitive flexibility. Participants match symbols to numbers based on a provided key, within a time limit. The score is based on the number of correct matches, with faster and more accurate responses indicating better cognitive function [53].

### 4.5 Patient-reported outcome measures

Fatigue was measured using the validated 8-item Checklist Individual Strength Revised (CIS8R), which evaluates subjective fatigue based on eight questions regarding fatigue intensity over the past two weeks [54, 55]. A higher score indicates greater fatigue. Sleep quality was assessed using the 19-item Pittsburgh Sleep Quality Index (PSQI), which measures sleep quality and disturbances across seven domains [56, 57]. A higher score indicated poorer sleep quality. Health-related quality of life (HRQoL) was assessed using the commonly applied Short Form 12 (SF12), a reliable and validated 12-item instrument, resulting in 8 subscales, which can be combined into a physical and mental component score [58–60]. A higher score indicates better HRQoL.

### 4.6 Demographic, Clinical, and Laboratory Variables

Demographic and transplant-related data were collected by questionnaires or extracted from the patient’s medical records. Sex was defined as sex assigned at birth. For commonly performed laboratory measurements in clinical care (i.e., leukocytes, estimated glomerular filtration rate, albumin, hemoglobin, sodium, creatinine, glucose, urea) the most recent result was extracted from the patient’s medical records. For the other laboratory measurements, which are less frequently assessed in clinical care, results from the most recent TransplantLines study visit were used to minimize indication bias. For recipients, median time between data extraction from the medical file and the SGMA was 0.12 years (*IQR* = 0.03 − 0.44); for lab results from the latest study visit, 2.73 years (*IQR* = 0.74 − 6.09). For donors, the median time between medical file data and the SGMA was 0.74 years (*IQR* = 0.36 − 1.78); for study visit lab results, 2.65 years (*IQR* = 1.22 − 4.08). Laboratory measurements were performed using routine laboratory methods. eGFR was calculated using the Chronic Kidney Disease Epidemiology Collaboration equation [61]. Iron deficiency was defined using both ferritin and transferrin saturation levels. For kidney donors, iron deficiency was defined as ferritin levels below 30 *g*/*L* in males and below 15 *g*/*L* in females. For recipients, iron deficiency was defined by a combination of ferritin levels below 100 *g*/*L* and transferrin saturation below 20% [62, 63]. Individuals not meeting these criteria were classified as not iron deficient. Medication use was extracted from the patient’s medical records.

### 4.7 Statistical analyses

For all sections, data preprocessing and statistical analyses were conducted with R [v.4.3.1, 64]. All data was visualized using ggplot2 [v.3.5.1, 65]. Regression analyses were conducted using *lme4* [v.1.1-34, 66] and *lmerTest* [v.3.1-3 67] packages.

To generate the results reported in section 2.5, we fitted four variations of penalized regression (LASSO) models using R package *glmnet* [v.4.1-8, 68] to predict (a) the SGMA score for recipients, (b) the SGMA score for recipients adjusted for demographics age, sex and education level, (c) the SGMA score for donors, and (d) the SGMA score adjusted for demographics age, sex, and education level using all available blood markers in the TransplantLines Biobank. If necessary (skewness > 2 or < −2) lab markers were converted to normal distributions using a log-transform. All lab variables were standardized and adjusted for sex. Lab measurements were included if they contained less than 20% missing data in the sample of participants.

The LASSO model was fitted using 100-fold cross-validation to choose the most optimal elastic net hyper-parameters for mixture (alpha) and penalty (lambda). We report standardized LASSO regression coefficients and R-squared values for all four models.

## Data Availability

SGMA data and analysis scripts are publicly available. Due to patient confidentiality, the clinical data associated with the SGMA dataset are not publicly available but can be made available upon request. Access to this clinical dataset requires a minimal access procedure consisting of a request by email. A response will be provided within 2 weeks. This access procedure is to ensure that the clinical data are being requested for research/scientific purposes only and thus complies with the informed consent signed by TransplantLines participants, which specifies that the collected data will not be used by commercial parties.

https://osf.io/p7aus

## Declarations

## 4.8 Acknowledgments

The authors would like to thank Dr. Michele F. Eisinga for his contributions to interpretation of iron- and anemia-related blood markers and their relation to cognitive performance in this study.

The TransplantLines Biobank and Cohort study was supported by grants from Astellas BV (project code: TransplantLines Biobank and Cohort study); Chiesi Pharmaceuticals BV (project code: PA-SP/PRJ-2020-9136); and NWO/TTW via a partnership program with DSM, Animal Nutrition and Health, The Netherlands (project code: 14939). The project was co-financed by the Dutch Ministry of Economic Affairs and Climate Policy by means of so-called PPP-allowances, made available by the Top Sector Life Sciences & Health to stimulate public-private partnerships (project code: PPP-2019-032, PPP-2022-015). The funders had no role in the study design, data collection, analysis, reporting, or the decision to submit for publication.

## 4.9 Author contributions

Conceptualization, Methodology, Formal analyses: TJW, TJK, HvR, SJLB; Software: TJW, TJK; Resources: TJW, HvR, HH, AS (SGMA items and algorithm), TransplantLines Investigators,(TransplantLines Biobank), Writing– original draft: TJW, TJK; Writing–Review & Editing: all authors; Visualization: TW, Supervision: HvR, SJLB.

## 4.11 Materials and correspondence

Correspondence may be addressed to prof. dr. Hedderik van Rijn, MemoryLab Health BV, Stationsstraat 10, 9711AS, Groningen, The Netherlands via.

SGMA data and analysis scripts are publicly available at https://osf.io/p7aus/. Due to patient confidentiality, the clinical data associated with the SGMA dataset are not publicly available but can be made available upon request. Access to this clinical dataset requires a minimal access procedure consisting of a request by email. A response will be provided within 2 weeks. This access procedure is to ensure that the clinical data are being requested for research/scientific purposes only and thus complies with the informed consent signed by TransplantLines participants, which specifies that the collected data will not be used by commercial parties.

## 4.12 TransplantLines Investigators

Coby Annema, Stefan P. Berger, Hans Blokzijl, Frank A.J.A. Bodewes, Marieke T. de Boer, Kevin Damman, Martin H. de Borst, Isabelle J.C. Dielwart, Arjan Diepstra, Gerard Dijkstra, Rianne M. Douwes, Caecilia S.E. Doorenbos, Michele F. Eisenga, Michiel E. Erasmus, C. Tji Gan, Antonio W. Gomes-Neto, Eelko Hak, Bouke G. Hepkema, Marius C. van den Heuvel, Jip Jonker, Frank Klont, Tim J. Knobbe, Daan Kremer, Coretta van Leer-Buter, Henri G.D. Leuvenink, Marco van Londen, Willem S. Lexmond, Vincent E. de Meijer, Hubert G.M. Niesters, Gertrude J. Nieuwenhuis-Moeke, L. Joost van Pelt, Robert A. Pol, Anna M. Posthumus, Adelita V. Ranchor, Jan-Stephan F. Sanders, Marion J. Siebelink, Riemer H.J.A. Slart, J. Cas Swarte, Daan J. Touw, Charlotte A. te Velde-Keyzer, Erik A.M. Verschuuren, Michel J. Vos, Rinse K. Weersma, Stephan J.L. Bakker

## 5 Supplementary materials

### Flow diagram

**Supplementary Figure 1.**
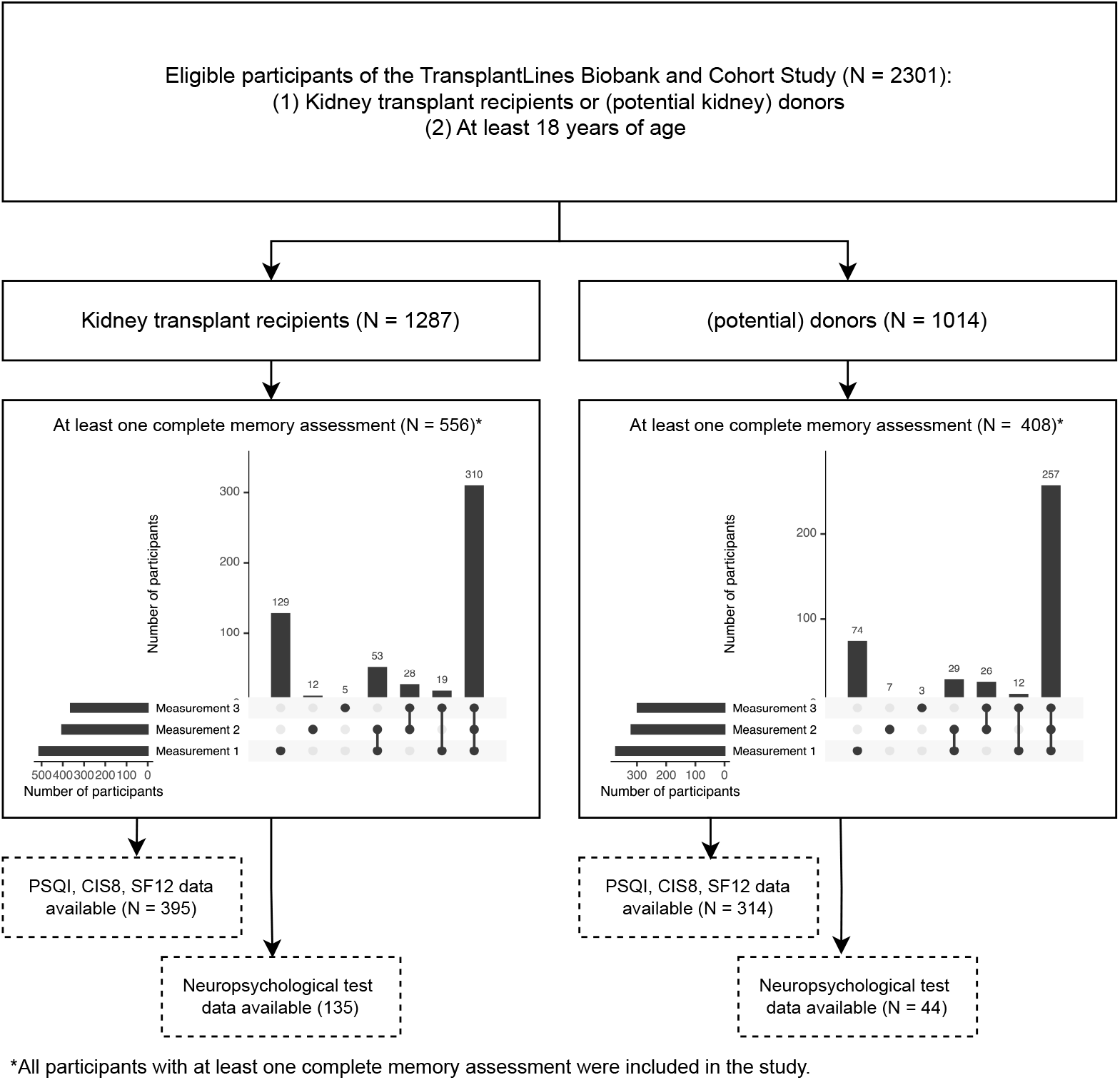
Flow diagram for participant inclusion, including upset plots for the three memory measurements in this study.

### Study Timeline

**Supplementary Figure 2.**
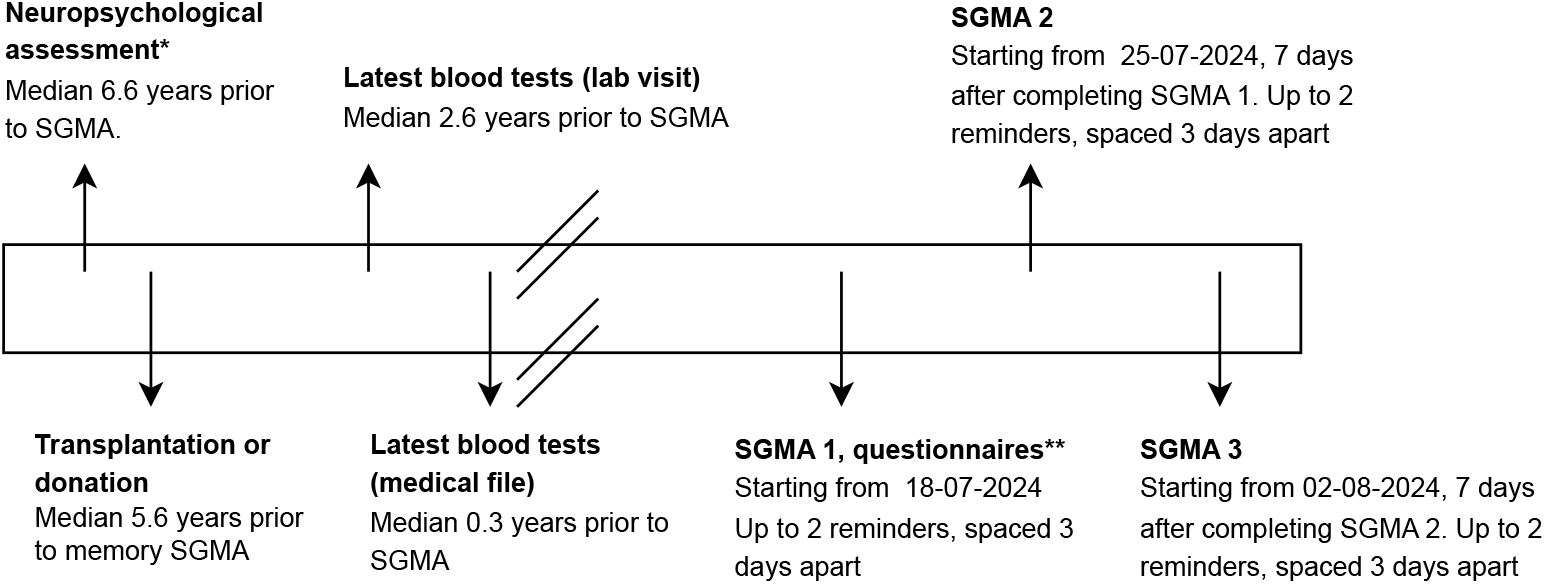
Timeline for key study time points. *Neuropsychological assessments were completed exclusively between 2015–2021. **Questionnaires included basic demographics, patient-reported outcome measures (CIS8R, SF12, PSQI), and ratings of subjective memory score.

### Sample characteristics for responders and non-responders

**Supplementary Table 1.** Sample characteristics for non-responders and responders. For time since donation or transplantation the median (IQR) value is shown, the other rows show means (SD) or absolute numbers and proportions.

|  | **Not responded** | **Responded** | *p* |
| --- | --- | --- | --- |
| **Recipients** |  |  |  |
| N | 731 | 556 |  |
| Age (years) | 58.37 (14.28) | 58.59 (12.64) | 0.77 |
| N female | 263 (36%) | 222 (40%) | 0.18 |
| Years since transplantation | 6.40 (2.50–11.32) | 6.06 (3.10–10.94) | 0.94 |
| N pre-transplant dialysis | 446 (61%) | 311 (56%) | 0.08 |
| N living donor | 402 (55%) | 344 (62%) | **0.01** |
| eGFR* (mL/min/1.73 m<sup>2</sup>) | 52.47 (18.10) | 53.16 (17.53) | 0.50 |
| **Donors/potential donors** |  |  |  |
| N | 606 | 408 |  |
| Age (years) | 60.58 (12.21) | 63.27 (10.77) | < **0.001** |
| N female | 303 (50%) | 228 (56%) | 0.09 |
| Years since donation | 5.76 (2.76–7.83) | 4.98 (2.21–7.72) | 0.26 |
| eGFR* (mL/min/1.73 m<sup>2</sup>) | 68.57 (16.92) | 63.09 (13.80) | < **0.001** |

\*estimated glomerular filtration rate (creatinine).

### Neuropsychological tests for recipients and donors

**Supplementary Table 2.** Means and standard deviations of neuropsychological tests for recipients and donors.

| Test | Recipient | Donor | *p* | Domain |
| --- | --- | --- | --- | --- |
| Trail Making Test-B | 66.85 (25.62) | 60.42 (21.12) | 0.10 | Attention |
| Clock Drawing | 11.89 (1.80) | 11.91 (1.41) | 0.94 | Executive Function |
| Dutch Reading Test | 13.79 (0.42) | 13.88 (0.27) | 0.10 | Language |
| Word fluency (animals) | 24.02 (5.53) | 28.07 (5.36) | < **0.001** | Language |
| Word fluency (letters) | 35.26 (12.11) | 42.95 (11.40) | < **0.001** | Language |
| Word fluency (professions) | 16.68 (3.97) | 20.42 (4.58) | < **0.001** | Language |
| 15 Words Test - Delayed retention | 9.58 (2.96) | 9.17 (3.19) | 0.47 | Memory |
| 15 Words Test - Immediate recall | 43.44 (9.51) | 42.41 (10.73) | 0.59 | Memory |
| Digit Span Backwards | 8.69 (1.86) | 9.14 (1.98) | 0.19 | Memory |
| Digit Span Forwards | 7.88 (2.13) | 8.14 (1.76) | 0.43 | Memory |
| Symbol Digits Modalities | 49.81 (10.01) | 50.67 (8.43) | 0.58 | Processing Speed |
| Trail Making Test-A | 30.88 (11.50) | 30.03 (10.72) | 0.66 | Processing Speed |

### Time since neuropsychological assessment

**Supplementary Figure 3.**
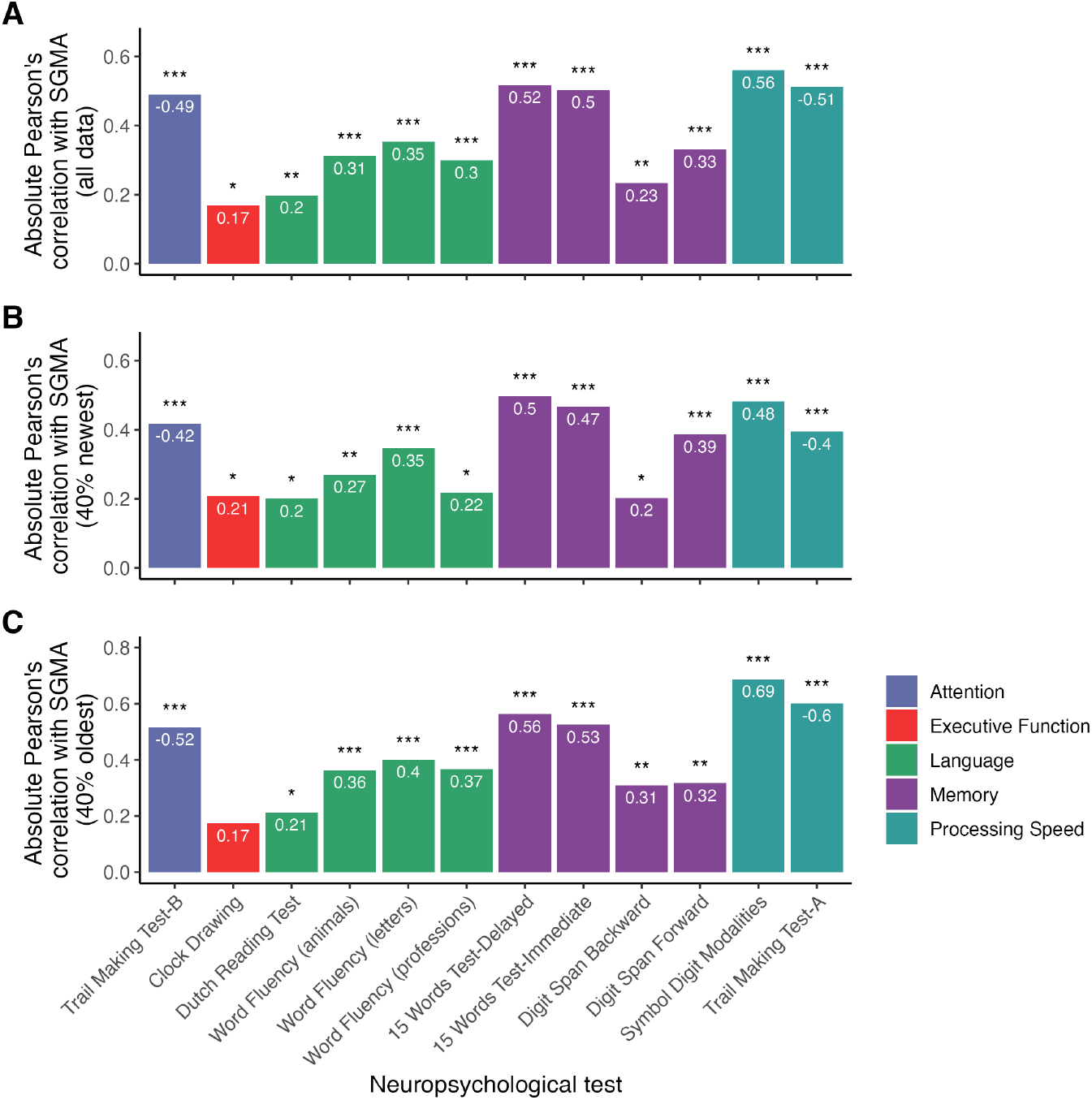
**Panel A** shows absolute Pearson’s correlations between SGMA scores and a range of neuropsychological tests across the full dataset (corresponding to main Figure 3). **Panel B** shows correlations restricted to participants with the most recent neuropsychological assessment (40% most recent observations). **Panel C** shows correlations restricted to participants with the oldest assessments (40% oldest observations). Bars are color-coded by cognitive domain. * * *: *p* < 0.001; **: *p* < 0.01; *: *p* < 0.05.

### Subjective memory ratings

**Supplementary Figure 4.**
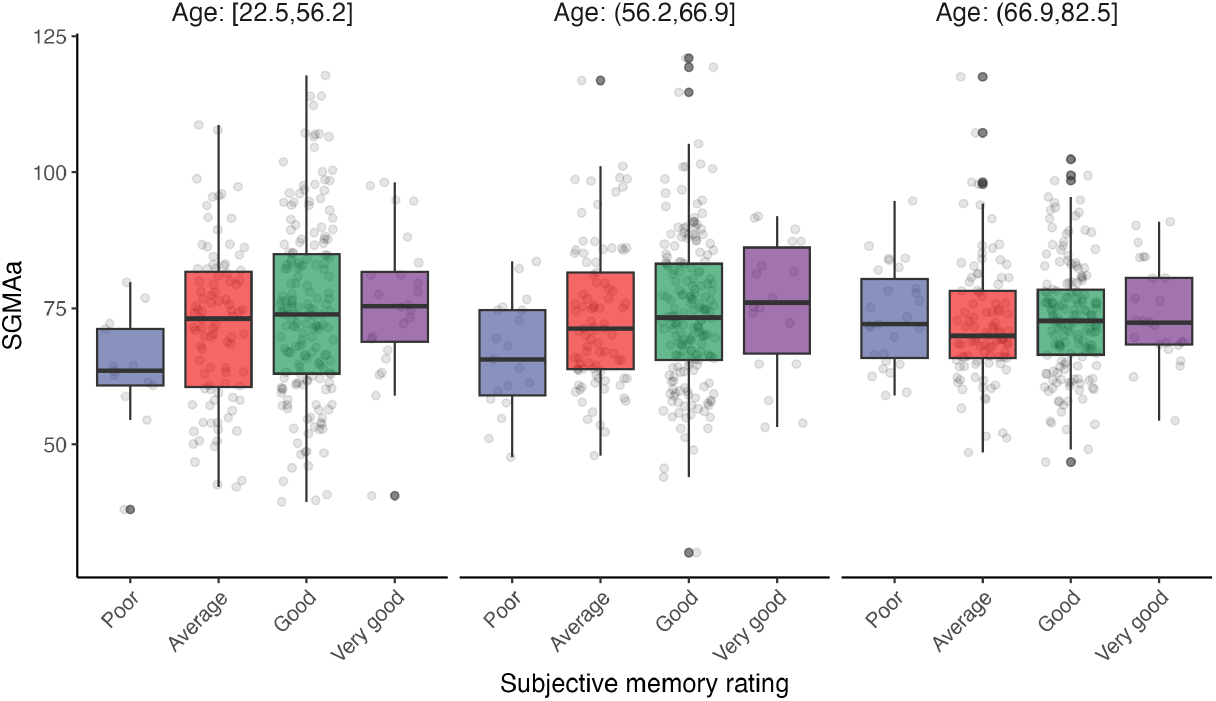
Relationship between objective memory performance as measured by the SGMA, adjusted for demographics, and subjective ratings of memory performance. Participants are divided into three equally-sized age brackets: between 22.5–56.2 years, between 56.2–66.9 years and between 66.9–82.5 years. While in the younger and middle-age participants, there was an association between subjective and objective memory, this relationship was not found in elderly participants.

### Sensitivity analysis

**Supplementary Figure 5.**
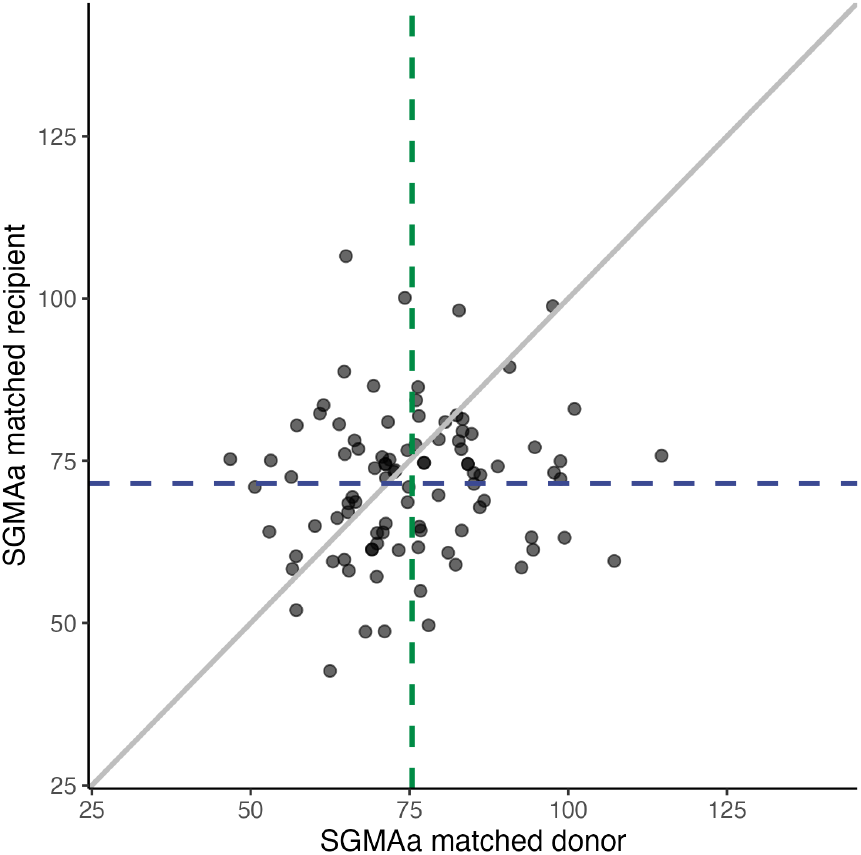
Scatterplot of SGMAa scores in KTR versus SGMAa scores of their donors. Each dot represents one matched pair. The solid diagonal line indicates equal performance between groups, while dashed lines show mean SGMAa scores for donors (green) and recipients (blue). Even under matched conditions, recipients scored lower on SGMAa than donors, confirming reduced memory performance in KTR.

### Primary kidney disease

**Supplementary Table 3.** Numbers indicate p-values for pairwise comparisons of SGMAa scores of KTR with etiologies of kidney failure.

|  | Cystic<br>KD | Glomerulo-<br>nephritis | Interstitial-nephritis | Other congenital<br>and hereditary KD | Renal vascular disease,<br>excluding vasculitis | Diabetes<br>Mellitus | Other multisystem<br>diseases | Unknown |
| --- | --- | --- | --- | --- | --- | --- | --- | --- |
| Glomerulonephritis | 0.09 |  |  |  |  |  |  |  |
| Interstitial-nephritis | 0.07 | 0.53 |  |  |  |  |  |  |
| Other congenital and hereditary KD | 0.62 | 0.66 | 0.41 |  |  |  |  |  |
| Renal vascular disease, excluding vasculitis | 0.78 | 0.32 | 0.19 | 0.80 |  |  |  |  |
| Diabetes Mellitus | **0.03** | 0.24 | 0.59 | 0.22 | 0.08 |  |  |  |
| Other multisystem diseases | 0.38 | 0.75 | 0.44 | 0.87 | 0.61 | 0.21 |  |  |
| Unknown | 0.11 | 0.88 | 0.65 | 0.61 | 0.31 | 0.32 | 0.68 |  |
| Other | 0.44 | 0.96 | 0.67 | 0.78 | 0.58 | 0.40 | 0.87 | 0.90 |

**Supplementary Figure 6.**
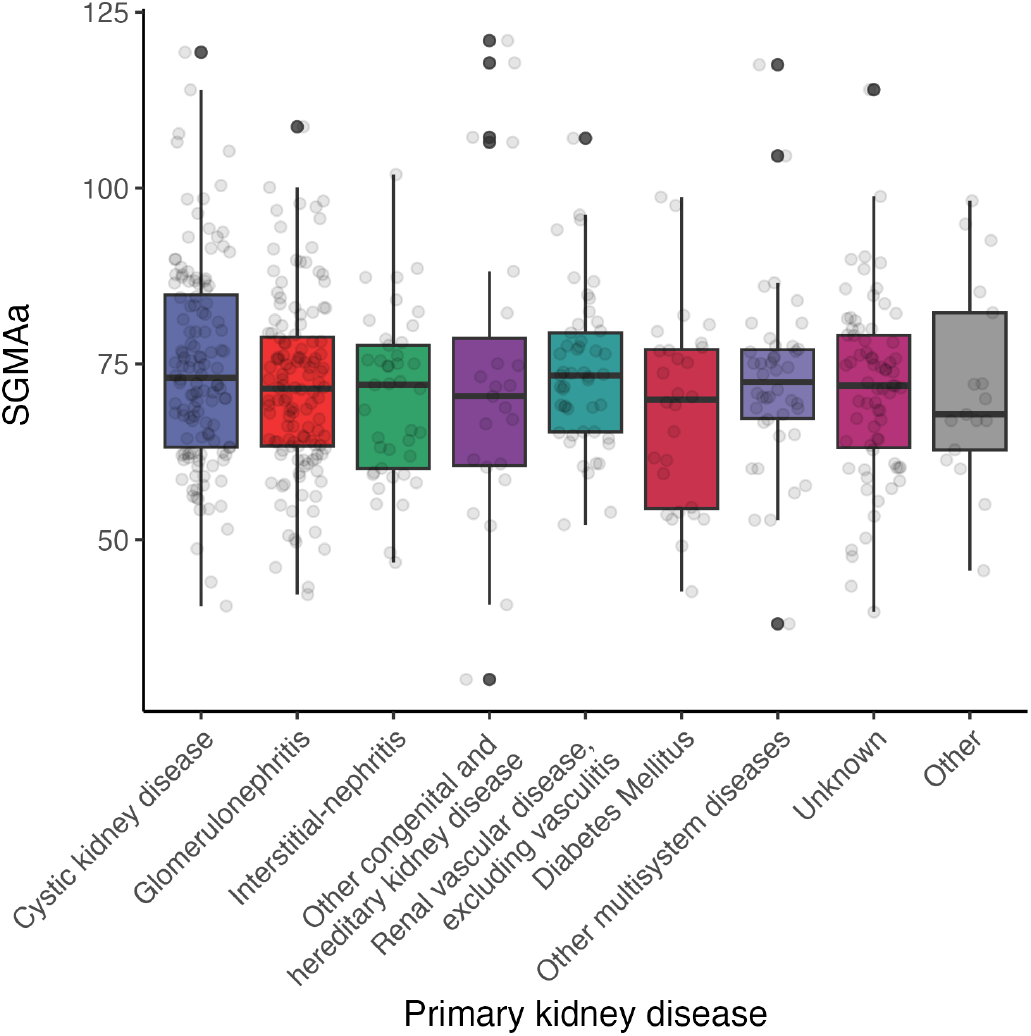
SGMAa values for KTR with different primary kidney disease etiologies.

### Lab marker variance

**Supplementary Figure 7.**
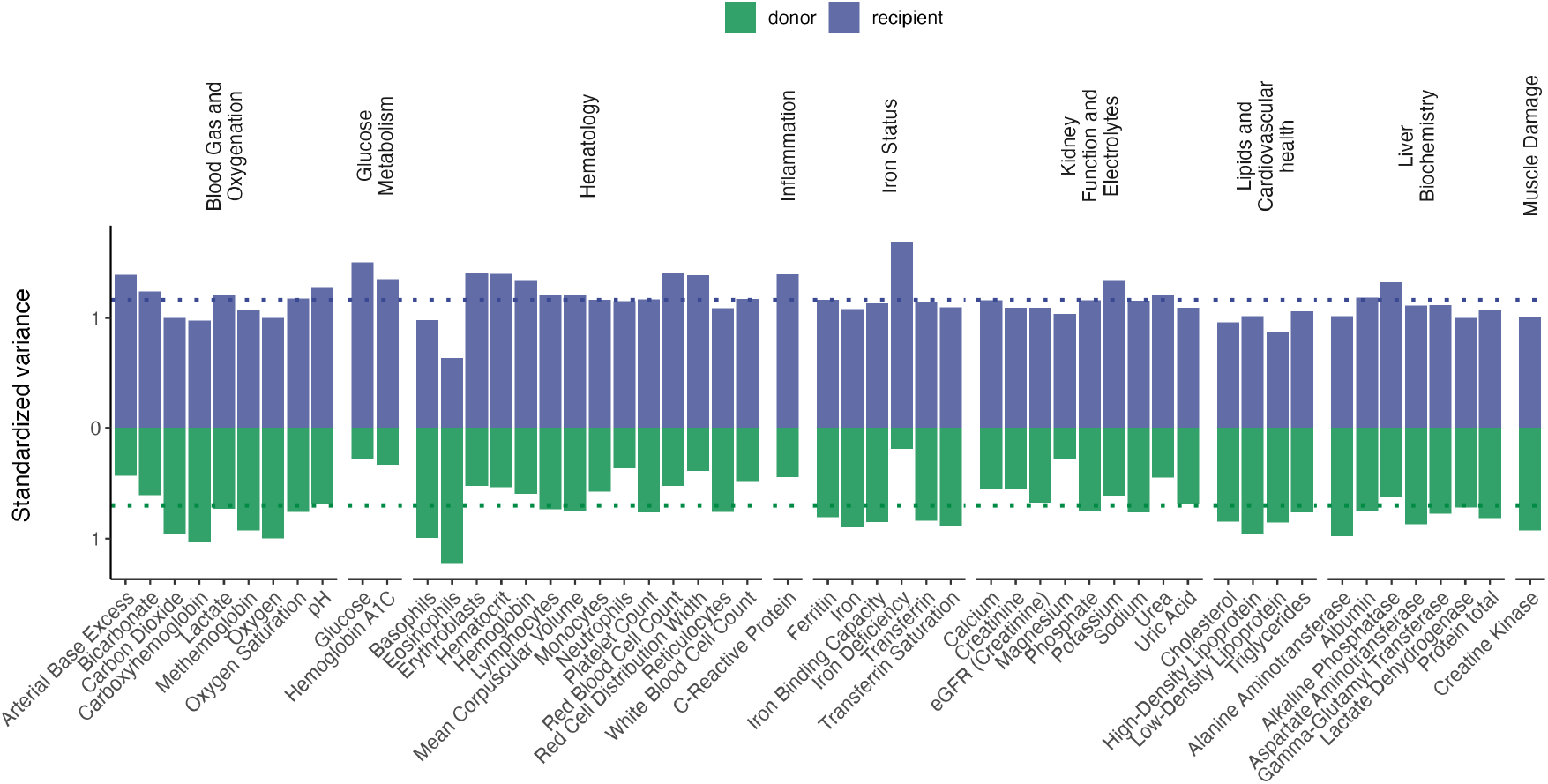
Average standardized variance by lab measurement, separated for recipients (blue, plotted upwards) and (potential) donors (green, plotted downwards). Dotted lines show averages across markers.

### Blood marker correlations

**Supplementary Table 4.** Correlation coefficients for the association between lab values and memory score, stratified by category and outcome measure. Values between brackets indicate p-values.

| Blood marker | Category | SGMA, all | SGMAa, all | SGMA, recipients | SGMAa, recipients | SGMA, donors | SGMAa, donors |
| --- | --- | --- | --- | --- | --- | --- | --- |
| 1 Arterial Base Excess | Blood Gas and Oxygenation | -0.01 (0.878) | 0.06 (0.087) | -0.01 (0.892) | 0.04 (0.344) | 0.01 (0.835) | 0.05 (0.347) |
| 2 Bicarbonate | Blood Gas and Oxygenation | 0 (0.990) | 0.06 (0.090) | -0.02 (0.706) | 0.05 (0.334) | 0.04 (0.387) | 0.04 (0.406) |
| 3 Carbon Dioxide | Blood Gas and Oxygenation | 0.02 (0.504) | 0.05 (0.176) | -0.01 (0.843) | 0.04 (0.447) | 0.07 (0.150) | 0.04 (0.436) |
| 4 Carboxyhemoglobin | Blood Gas and Oxygenation | 0.03 (0.396) | -0.02 (0.591) | 0.05 (0.324) | 0.01 (0.846) | 0.01 (0.869) | -0.06 (0.273) |
| 5 Lactate | Blood Gas and Oxygenation | -0.08 (0.016) | -0.06 (0.074) | -0.09 (0.049) | -0.07 (0.132) | -0.07 (0.140) | -0.02 (0.630) |
| 6 Methemoglobin | Blood Gas and Oxygenation | 0.03 (0.422) | 0 (0.896) | 0 (0.976) | 0 (0.950) | 0.06 (0.215) | 0.02 (0.673) |
| 7 Oxygen | Blood Gas and Oxygenation | -0.02 (0.577) | -0.01 (0.699) | 0.02 (0.636) | 0.01 (0.891) | -0.07 (0.153) | -0.04 (0.387) |
| 8 Oxygen Saturation | Blood Gas and Oxygenation | -0.02 (0.551) | -0.01 (0.726) | 0.01 (0.802) | 0 (0.953) | -0.07 (0.183) | -0.05 (0.313) |
| 9 pH | Blood Gas and Oxygenation | -0.03 (0.332) | 0 (0.963) | 0 (0.945) | 0 (0.958) | -0.08 (0.096) | -0.01 (0.804) |
| 10 Glucose | Glucose Metabolism | -0.11 (0.001) | -0.05 (0.152) | -0.09 (0.039) | -0.01 (0.872) | -0.21 (0.000) | -0.10 (0.041) |
| 11 Hemoglobin A1C | Glucose Metabolism | -0.17 (0.000) | -0.06 (0.067) | -0.20 (0.000) | -0.05 (0.271) | -0.17 (0.001) | 0 (0.987) |
| 12 Basophils | Hematology | -0.02 (0.491) | -0.02 (0.598) | -0.04 (0.440) | -0.06 (0.192) | -0.01 (0.885) | 0.01 (0.777) |
| 13 Eosinophils | Hematology | 0.02 (0.627) | 0.04 (0.270) | 0.03 (0.551) | -0.01 (0.825) | 0.01 (0.845) | 0.04 (0.422) |
| 14 Erythroblasts | Hematology | -0.04 (0.259) | -0.04 (0.274) | -0.03 (0.501) | -0.04 (0.379) | -0.06 (0.258) | -0.01 (0.905) |
| 15 Hematocrit | Hematology | -0.07 (0.029) | 0.01 (0.692) | -0.09 (0.057) | 0.02 (0.639) | -0.05 (0.325) | 0 (0.973) |
| 16 Hemoglobin | Hematology | -0.05 (0.105) | 0.02 (0.482) | -0.06 (0.188) | 0.03 (0.558) | -0.04 (0.370) | 0 (0.949) |
| 17 Lymphocytes | Hematology | 0.02 (0.520) | -0.02 (0.604) | 0.05 (0.304) | -0.04 (0.453) | -0.02 (0.689) | -0.02 (0.738) |
| 18 Mean Corpuscular Volume | Hematology | -0.04 (0.197) | -0.03 (0.363) | -0.02 (0.622) | -0.03 (0.52) | -0.08 (0.118) | -0.04 (0.379) |
| 19 Monocytes | Hematology | -0.08 (0.018) | -0.08 (0.019) | -0.11 (0.020) | -0.09 (0.044) | -0.05 (0.350) | 0 (0.963) |
| 20 Neutrophils | Hematology | -0.10 (0.004) | -0.14 (0.000) | -0.16 (0.001) | -0.16 (0.001) | -0.02 (0.635) | -0.04 (0.431) |
| 21 Platelet Count | Hematology | 0.01 (0.665) | -0.03 (0.329) | 0.01 (0.743) | -0.07 (0.133) | 0.02 (0.745) | -0.01 (0.882) |
| 22 Red Blood Cell Count | Hematology | -0.04 (0.221) | 0.03 (0.336) | -0.06 (0.168) | 0.04 (0.441) | 0 (0.951) | 0.04 (0.435) |
| 23 Red Cell Distribution Width | Hematology | -0.05 (0.145) | -0.02 (0.582) | -0.07 (0.111) | -0.01 (0.879) | -0.02 (0.750) | 0.03 (0.590) |
| 24 Reticulocytes | Hematology | -0.10 (0.002) | -0.12 (0.000) | -0.14 (0.002) | -0.16 (0.001) | -0.06 (0.208) | -0.03 (0.513) |
| 25 White Blood Cell Count | Hematology | -0.08 (0.015) | -0.13 (0.000) | -0.12 (0.008) | -0.15 (0.001) | -0.04 (0.478) | -0.03 (0.519) |
| 26 C-Reactive Protein | Inflammation | -0.09 (0.007) | -0.07 (0.039) | -0.07 (0.128) | -0.04 (0.348) | -0.16 (0.001) | -0.09 (0.083) |
| 27 Ferritin | Iron Status | -0.06 (0.089) | 0.01 (0.752) | 0.06 (0.199) | 0.10 (0.026) | -0.24 (0.000) | -0.14 (0.006) |
| 28 Iron | Iron Status | 0.02 (0.540) | 0.05 (0.148) | 0.10 (0.029) | 0.08 (0.066) | -0.09 (0.063) | -0.02 (0.725) |
| 29 Iron Binding Capacity | Iron Status | 0 (0.996) | -0.01 (0.737) | -0.05 (0.272) | -0.09 (0.041) | 0.08 (0.128) | 0.10 (0.045) |
| 30 Iron Deficiency | Iron Status | -0.05 (0.210) | -0.10 (0.007) | -0.11 (0.047) | -0.14 (0.013) | 0.01 (0.036) | 0.06 (0.197) |
| 31 Transferrin | Iron Status | 0.01 (0.853) | 0 (0.959) | -0.04 (0.422) | -0.07 (0.127) | 0.07 (0.147) | 0.10 (0.041) |
| 32 Transferrin Saturation | Iron Status | 0.01 (0.737) | 0.04 (0.269) | 0.10 (0.028) | 0.10 (0.034) | -0.12 (0.018) | -0.06 (0.261) |
| 33 Calcium | Kidney Function and Electrolytes | -0.03 (0.384) | 0.02 (0.485) | -0.03 (0.528) | 0.05 (0.259) | -0.04 (0.404) | 0.07 (0.179) |
| 34 Creatinine | Kidney Function and Electrolytes | -0.01 (0.671) | -0.08 (0.013) | 0 (0.944) | -0.06 (0.158) | -0.05 (0.268) | -0.04 (0.418) |
| 35 Magnesium | Kidney Function and Electrolytes | -0.04 (0.210) | 0.02 (0.487) | -0.02 (0.594) | -0.01 (0.857) | -0.11 (0.033) | -0.08 (0.113) |
| 36 Phosphate | Kidney Function and Electrolytes | 0 (0.940) | 0.02 (0.652) | -0.01 (0.761) | -0.03 (0.468) | 0.03 (0.52) | 0.05 (0.269) |
| 37 Potassium | Kidney Function and Electrolytes | -0.02 (0.598) | 0 (0.895) | -0.04 (0.396) | -0.02 (0.684) | 0.02 (0.647) | 0.05 (0.268) |
| 38 Sodium | Kidney Function and Electrolytes | 0.02 (0.626) | 0 (0.958) | 0.06 (0.170) | 0.04 (0.379) | -0.07 (0.181) | -0.04 (0.478) |
| 39 Urea | Kidney Function and Electrolytes | -0.08 (0.014) | -0.06 (0.096) | -0.08 (0.096) | -0.05 (0.313) | -0.13 (0.008) | 0.01 (0.776) |
| 40 Uric Acid | Kidney Function and Electrolytes | -0.07 (0.038) | -0.06 (0.063) | -0.07 (0.135) | -0.08 (0.079) | -0.09 (0.070) | 0.02 (0.625) |
| 41 eGFR (Creatinine) | Kidney Function and Electrolytes | 0.14 (0.000) | 0.09 (0.008) | 0.11 (0.008) | 0.07 (0.085) | 0.23 (0.000) | 0.05 (0.339) |
| 42 Cholesterol | Lipids and Cardiovascular health | -0.04 (0.218) | 0.04 (0.209) | -0.01 (0.748) | 0.04 (0.369) | -0.08 (0.105) | -0.02 (0.745) |
| 43 High-Density Lipoprotein | Lipids and Cardiovascular health | 0.03 (0.378) | 0.06 (0.088) | 0.02 (0.721) | 0.03 (0.567) | 0.05 (0.317) | 0.08 (0.124) |
| 44 Low-Density Lipoprotein | Lipids and Cardiovascular health | -0.04 (0.270) | 0.04 (0.216) | 0 (0.937) | 0.04 (0.333) | -0.08 (0.091) | -0.03 (0.518) |
| 45 Triglycerides | Lipids and Cardiovascular health | -0.06 (0.075) | -0.05 (0.123) | -0.05 (0.296) | -0.02 (0.675) | -0.09 (0.075) | -0.05 (0.357) |
| 46 Alanine Aminotransferase | Liver Biochemistry | -0.01 (0.838) | 0.02 (0.482) | 0.03 (0.488) | 0.05 (0.244) | -0.06 (0.244) | -0.03 (0.559) |
| 47 Albumin | Liver Biochemistry | 0.07 (0.039) | 0.02 (0.609) | 0.09 (0.034) | 0.02 (0.673) | 0.03 (0.601) | 0 (0.983) |
| 48 Alkaline Phosphatase | Liver Biochemistry | -0.03 (0.404) | -0.01 (0.778) | 0 (0.925) | 0 (1) | -0.09 (0.064) | -0.02 (0.749) |
| 49 Aspartate Aminotransferase | Liver Biochemistry | -0.03 (0.386) | 0.01 (0.711) | 0.01 (0.771) | 0.05 (0.279) | -0.09 (0.063) | -0.05 (0.340) |
| 50 Gamma-Glutamyl Transferase | Liver Biochemistry | -0.07 (0.033) | 0.01 (0.874) | -0.06 (0.195) | 0.03 (0.529) | -0.10 (0.047) | 0.02 (0.762) |
| 51 Lactate Dehydrogenase | Liver Biochemistry | -0.12 (0.001) | -0.05 (0.111) | -0.13 (0.004) | -0.01 (0.763) | -0.12 (0.017) | -0.04 (0.385) |
| 52 Protein total | Liver Biochemistry | -0.04 (0.240) | -0.04 (0.266) | 0.02 (0.728) | -0.04 (0.369) | -0.12 (0.014) | -0.08 (0.108) |
| 53 Creatine Kinase | Muscle Damage | -0.01 (0.658) | -0.02 (0.492) | -0.01 (0.771) | -0.05 (0.307) | -0.02 (0.736) | -0.03 (0.560) |

### Time since blood test

**Supplementary Figure 8.**
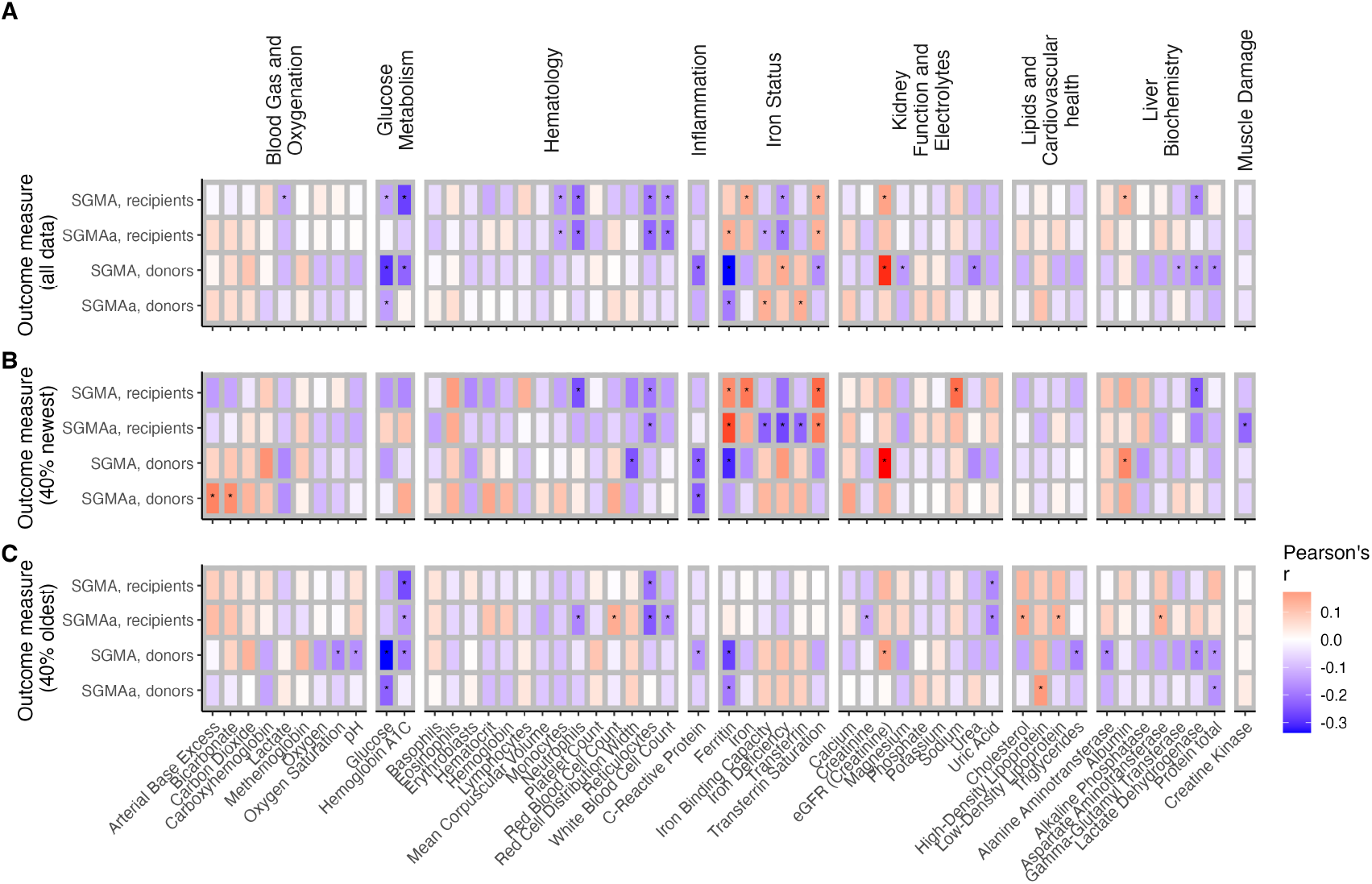
**Panel A** shows individual Pearson’s correlation coefficients between SGMA scores for recipients, SGMAa scores for recipients, SGMA scores for donors, and SGMAa scores for donors and blood measurements across the full dataset (corresponding to main Figure 5. **Panel B** shows correlations restricted to participants with the most recent blood marker data (40% most recent observations). **Panel C** shows correlations restricted to participants with the oldest assessments (40% oldest observations). Stars indicate significance at the level of *: *p* < 0.05.

## Footnotes

1 Note that Figure 4B shows the age-adjusted SGMA scores, and therefore no decline over time as a result of normal aging is shown. The flat lines in the figure show that there is no *steeper* decline in memory function post transplantation.

